# Feasibility, Acceptability, and Effectiveness of Non-Pharmaceutical Interventions for Infectious Disease Prevention and Control in Crisis-Affected Settings and Informal Settlements: A Scoping Review

**DOI:** 10.1101/2021.08.20.21262352

**Authors:** Jonathan A. Polonsky, Sangeeta Bhatia, Keith Fraser, Arran Hamlet, Janetta Skarp, Isaac J. Stopard, Stéphane Hugonnet, Laurent Kaiser, Christian Lengeler, Karl Blanchet, Paul Spiegel

## Abstract

**Introduction:** Non-pharmaceutical interventions (NPIs) are a crucial suite of measures to prevent and control infectious disease outbreaks. They are particularly important for crisis-affected populations that typically reside in settings characterised by overcrowding, inadequate access to healthcare and resource limitations. To describe the landscape of research and identify evidence gaps concerning the acceptability, feasibility, and effectiveness of NPIs among crisis-affected populations and informal settings, we conducted a systematic scoping review of the published evidence.

**Methods:** We systematically reviewed peer-reviewed articles published between 1970 and 2020 to collate available evidence on the feasibility, acceptability, and effectiveness of NPIs in crisis-affected populations and informal settlements. We performed quality assessments of each study using a standardised questionnaire.

**Results:** Our review included 158 studies published in 85 peer-reviewed articles. Most research used low quality study designs. The acceptability, feasibility, and effectiveness of NPIs was highly context dependent. In general, simple and cost-effective interventions such as community-level environmental cleaning and provision of water, sanitation and hygiene services, and distribution of items for personal protection such as insecticide-treated nets, were both highly feasible and acceptable. Logistical, financial, and human resource constraints affected both the implementation and sustainability of measures. Community engagement emerged as a strong factor contributing to the effectiveness of NPIs. Conversely, measures that involve potential restriction on personal liberty such as case isolation were found to be less acceptable to the community.

**Conclusion:** Overall, the evidence base was patchy, with substantial knowledge gaps between differing between settings and pathogens. Although implementation of NPIs presents unique practical challenges, it is critical that the lessons learned are shared with the wider community to build a robust evidence base.

**Summary box:** *What is already known?:*

- NPIs are a crucial suite of tools for the prevention and control of infectious diseases, either as a complement to, or in the absence of, effective pharmaceutical interventions (i.e., therapeutics and vaccination).
- Despite being disproportionately vulnerable, crisis-affected populations and those living in informal settlements are often neglected by research and guidance. NPIs are, however, vital and adaptation is necessary within these settings due to poor living conditions and resource limitations that intensify disease transmission risk.

*What are the new findings?:*

- We conducted a scoping review to produce a landscape analysis of the existing evidence concerning NPIs within crisis-affected settings and informal settlements.
- The existing evidence is patchy, uneven, occasionally contradictory, and of generally low quality, but building over time.
- Although limited, some findings are generalisable across settings, populations and NPIs.

*What do the new findings imply?:*

- There is a need for greater investments in research to strengthen the guidance and policies on NPIs in these settings.
- In particular, upstream pilot feasibility and acceptability studies should be conducted prior to the widespread roll-out of interventions to ensure they are feasible, acceptable, and ultimately effective for the target populations.

## Introduction

Non-pharmaceutical interventions (NPIs), also referred to as Public Health and Social Measures, are an important suite of health interventions used to reduce transmission and mitigate impacts of infectious diseases, particularly during early phases of epidemics when effective pharmaceutical interventions (i.e. therapeutics and vaccination) are not yet available or widely accessible [1].

NPIs range from lighter-touch personal protective measures, such as hand-hygiene and mask-wearing, to more stringent restrictions applied at community or societal levels, such as quarantines and travel bans, that are specifically geared to limiting person-to-person spread no matter the mode of transmission. Recently, there has been substantial research and guidance on implementing NPIs against pandemic influenza [2–6] and COVID-19 [7–13].

Humanitarian crises are characterised by limited access to resources, chronic underfunding, and poor accessibility. Within such settings, people are subject to an exacerbation of the inverse care law, whereby socially disadvantaged people have poorer access to more effective forms of disease prevention and control, specifically therapeutics and vaccinations, for which costs are higher and/or the global stockpile limited [14]. The number and geographic origin of crisis-affected individuals has expanded in recent years, with nearly 168 million people in need of humanitarian assistance and protection in 2020, an 86% increase since 2015 [15,16]. In addition, an estimated one billion people, or one-third of the world’s urban population, live in informal settlements (slums), which are characterised by similar constraints [17,18], and therefore also merit special consideration when planning disease prevention and control interventions.

While substantial research and guidance has been published exploring NPI implementation in non-emergency settings [1,19], little has been focussed on these millions of particularly vulnerable people. Furthermore, existing guidance does not account for context-specific risk factors that these settings present and that increase the risks of infectious disease outbreaks, including densely crowded and low-quality shelters, poor access to water, sanitation and hygiene (WASH) facilities, and nutritional stress [20,21]. Understanding the potential use and impact of NPIs in containing the spread of infectious diseases in these settings is, therefore, critical. Despite this, there remain important gaps in the understanding of three key areas that influence the extent to which implementation of NPIs will be successful: 1) feasibility (how realistic is it to implement the proposed interventions given logistical, financial, socio-cultural, and other barriers); 2) acceptability (how do affected communities perceive the interventions); and 3) effectiveness (do the measures impact disease transmission in real-life).

Therefore, we conducted a systematic scoping review of the published literature to better understand the range and quality of evidence on the feasibility, acceptability, and effectiveness of NPIs against infectious diseases among crisis-affected populations and in informal settlements. We present and synthesise this work, identifying gaps for further research to guide infectious disease prevention and control activities among these vulnerable populations.

## Materials and Methods

### Eligibility criteria

Articles describing research on NPIs against infectious diseases conducted in humanitarian crisis-affected settings and informal settlements (slums), two settings typified by similarly exacerbated infectious disease risks and resource constraints for disease prevention and control, were eligible for inclusion. We defined crisis-affected settings as those in which ‘an event or series of events has resulted in a critical threat to health, safety, security or well-being of a community or other large group of people’ [12], identifying five conditions, as previously described [22]: 1) progressive loss of livelihoods and deterioration of essential services due to ever-present risk of violence; 2) mass displacement into camp-like settlements; 3) displacement into neighbouring host communities; 4) sudden loss of livelihoods and rapid environmental change due to natural disaster, and 5) food crises. The United Nations Human Settlements Programme defines informal settlements as human settlements that have the following characteristics: inadequate access to safe water; inadequate access to sanitation and other infrastructure; poor structural quality of housing; overcrowding; and insecure residential status [17,23].

Articles written in English and published in peer-reviewed journals between 1970 and 2020 were eligible for inclusion. Both quantitative and qualitative studies were included, provided they contained relevant primary data.

These inclusion and exclusion criteria are summarised in greater detail in Table S1 (Supplementary Material).

### Search strategy and information sources

On the 24th of May 2020, we searched two bibliographic databases, PubMed and Web of Science, for entries dated between 1970-2020. The full search terms are listed in Supplementary Materials. This search was repeated on 5^th^ January 2021 to gather additional literature published after the initial search.

Studies were loaded into Covidence systematic review management software [24]. Duplicate entries were automatically detected and reduced to single entries. Review articles returned by the initial search were screened for potentially relevant references, which were added to the list of articles, with the reviews subsequently excluded. The following steps were then carried out on the deduplicated records independently by two reviewers. Irrelevant articles were excluded in two-steps; first by screening titles and abstracts and then by screening the full text of the remaining articles. Discrepancies and borderline cases were resolved through discussions between at least two reviewers.

### Data charting process

The final list of included articles was divided among reviewers for data charting (extraction and summary) using a structured questionnaire. The data items extracted related to several domains: 1) study metadata: authors, publication year, the years during which the NPIs were implemented, the region (using World Bank classification scheme) and country of study, country of origin of the crisis-affected population; 2) typologies of crisis, population, shelter, disease transmission mode, and level of intervention (personal protective, community, environmental, or disease control); 3) study descriptions: study design and size, research type (quantitative vs. qualitative), and measure (feasibility, accessibility or effectiveness); and 4) study detail: brief description of NPIs implemented and extraction of the main findings.

We charted information related to the following key outcomes and their adapted definitions [25,26]: 1) feasibility - the viability, practicability, or workability of the intervention. How possible or practicable it is to carry it out; 2) acceptability - a multi-faceted construct that reflects the extent to which people delivering or receiving a healthcare intervention consider it to be appropriate, based on anticipated or experienced cognitive and emotional responses to the intervention; 3) effectiveness - the extent to which a specific intervention, when deployed in the usual circumstances of practice, does what it is intended to do for a specified population. Where multiple research types and/or measures were reported in a single article (e.g., a quantitative finding on effectiveness and a qualitative finding on acceptability), data were entered across multiple rows.

### Critical appraisal of individual sources of evidence

The quality of the evidence of the included studies was assessed using the National Institutes of Health study quality assessment tool [27], with each rated “good”, “fair”, or “poor”. While this approach accounted for intra-design variability in quality, we also considered inter-design quality, using a rating scheme adapted from Yetley et al (2016) [28] that ranked study designs from highest to lowest, as follows: controlled intervention study, cohort study, case-control study, pre-post evaluation, cross-sectional study, and case study. These qualitative scores were taken into consideration at the data synthesis stage to help gauge the quality and strength of the evidence.

### Synthesis of results

We performed a qualitative synthesis of the findings, according to level of intervention (personal protective, community, environmental, or disease control), crisis type (conflict, natural disaster, complex emergency, food security crisis, and informal settlement,), and shelter type (camp or camp-like, non-displacement, hosted, and informal housing). Findings were grouped by the three key outcome measures (feasibility, acceptability, and effectiveness). We synthesised and summarised identifiable trends and commonalities in the findings and highlighted important gaps and limitations in the published research.

We report our findings according to the Preferred Reporting Items for Systematic Reviews and Meta-Analyses extension for scoping reviews (PRISMA-ScR) statement [29].

## Results

### Selection of sources of evidence

The database search initially identified 8399 studies, with a further 31 from review articles. After removing duplicates, 4518 (53.6%) remained, the majority of which (3972, 87.9%) were excluded at the abstract screening stage for not fitting the inclusion criteria, leaving 546 for full text screening. Of these, 461 (84.4%) were excluded: 242 did not concern an infectious disease or were not in a crisis-affected setting or informal settlement, 129 did not include an NPI, 62 were opinion pieces, the full text was not found for 18, six studies occurred before 1970, and four were unavailable in English), leaving 85 articles for inclusion [30–114] (Figure 1).

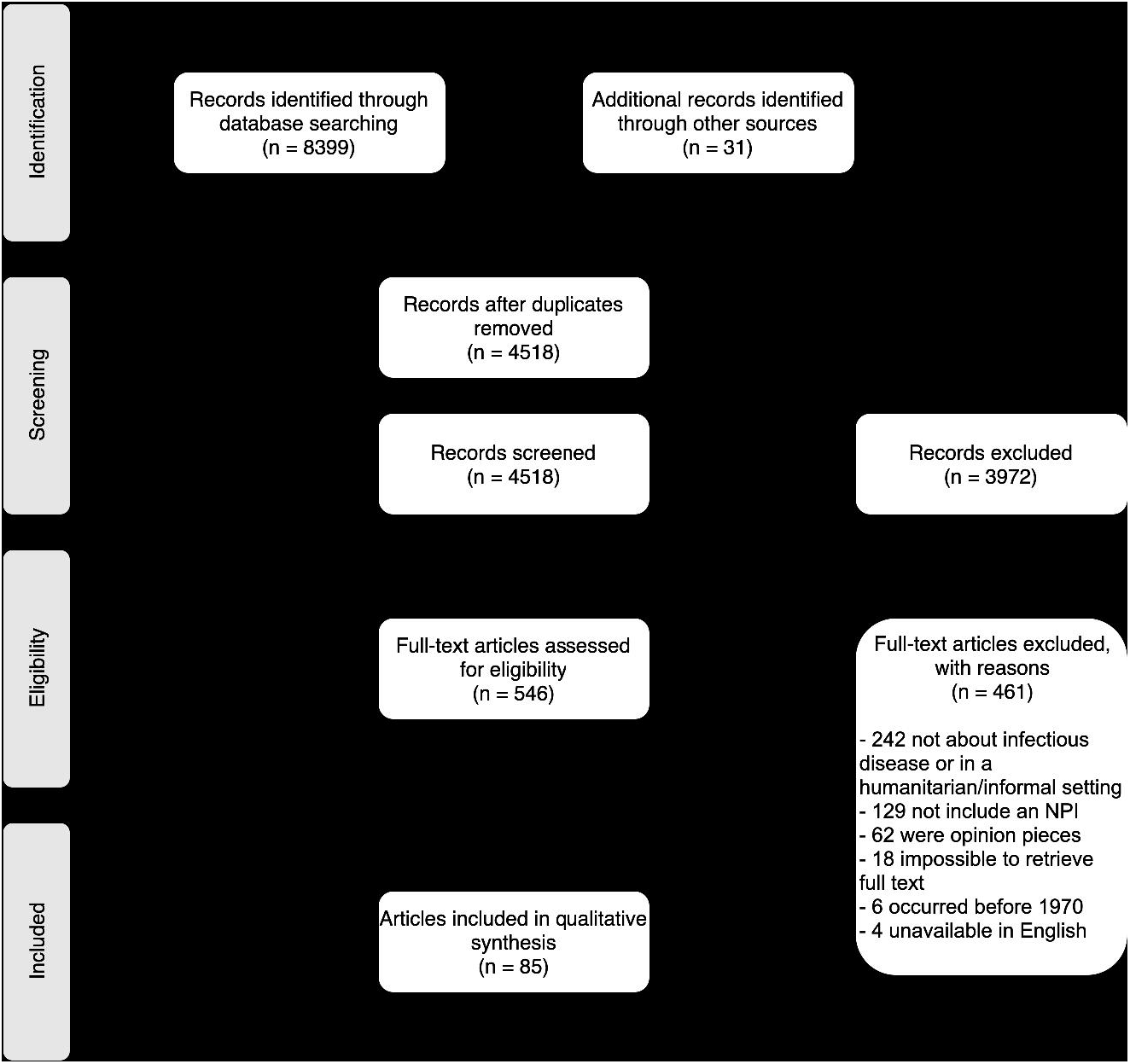

### Characteristics of sources of evidence

#### Characteristics of publications and setting

The volume of research increased over time, with no studies published prior to 1987, just two (2.4%) during the 1980s, eight (9.4%) during the 1990s, approximately one quarter (20, 23.5%) during the subsequent decade, and nearly two-thirds (55, 64.7%) published from 2010 to 2020 (Table 1).

**Table 1.** Key descriptive characteristics of articles included for data charting (N = 85).

| Characteristic | n (%) |
| --- | --- |
| ***Decade of publication*** |  |
| 1980s | 2 (2.4%) |
| 1990s | 8 (9.4%) |
| 2000s | 20 (23.5%) |
| 2010s | 55 (64.7%) |
| ***World Bank Region*** |  |
| Sub-Saharan Africa | 39 (45.9%) |
| South Asia | 17 (20.0%) |
| East Asia & Pacific | 13 (15.3%) |
| Latin America & Caribbean | 7 (8.2%) |
| North America | 4 (4.7%) |
| Middle East & North Africa | 4 (4.7%) |
| Europe & Central Asia | 1 (1.2%) |
| ***Crisis type*** |  |
| Conflict | 54 (63.5%) |
| Natural disaster | 19 (22.4%) |
| Informal housing | 10 (11.8%) |
| Complex emergency | 1 (1.2%) |
| Food security crisis | 1 (1.2%) |
| ***Shelter type*** |  |
| Camp or camp-like | 50 (58.8%) |
| Non-displacement | 14 (16.5%) |
| Informal housing | 11 (12.9%) |
| Not specified | 5 (5.9%) |
| Multiple | 4 (4.7%) |
| Hosted | 1 (1.2%) |
| ***Total*** | ***85 (100%)*** |

Studies were conducted in 38 countries across the seven World Bank regions, with the majority being in conflict-affected (N = 54, 63.5%) or natural disaster (N = 19, 22.4%) settings (Table 1). Nearly half (N = 39, 45.9%) of the articles were from the Sub-Saharan African region. The majority of these studies were in East and Central Africa, with nine from the Democratic Republic of the Congo (DRC) [36,49,61,64–66,72,105,107], seven from Sudan/South Sudan [30,39,79,80,104,106,108] and four from Uganda [75,76,78,99]. Almost all (N = 35, 89.7%) of the studies in this region were in conflict-affected settings, with two of the remaining four studies being the sole examples of both food crises [106] and complex emergencies [35] (Figure 2). South Asia (N = 17, 20.0%), and East Asia and the Pacific (N = 13, 15.3%), accounted for the bulk of the remainder, with fewer than ten studies in each of the remaining regions. Of these, four studies were conducted in the Middle East and North Africa, all in conflict-affected settings; three relating to the Syria conflict [32,41,52,110] and one during a cholera outbreak in Yemen [32]). There was just one study from Europe and Central Asia, during a Norovirus outbreak in a refugee camp in Germany [48]. The four studies in North America all took place in the United States of America (USA), all following natural disasters [43,50,58,63].

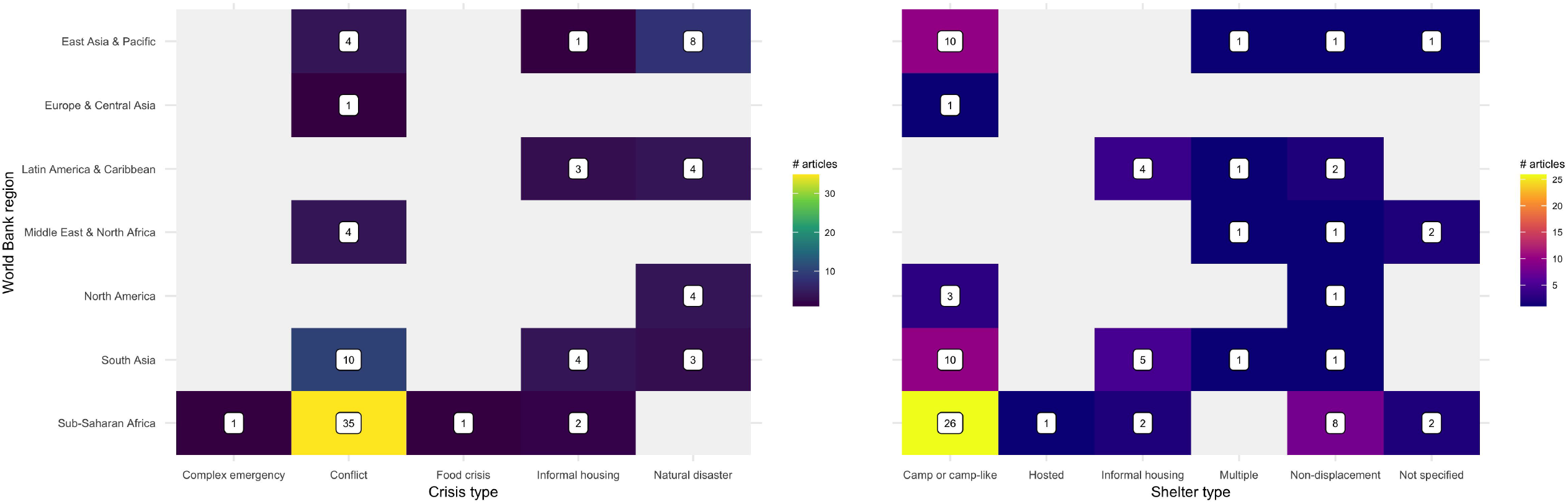

Over half of the studies (N = 50, 58.8%) were in camp or camp-like settings (Table 1, Figure 2), mostly (N = 29, 58.0%) among refugees but with a sizable proportion (N = 20, 40.0%) among internally displaced persons (IDPs). Most of the remaining studies were among crisis-affected but non-displaced (N = 16, 18.8%) and informally housed (i.e., slum dwelling) (N = 8, 9.4%) people. There were just two studies among hosted IDPs [92] and refugees [41] and one study among prisoners of war [100].

#### Characteristics of research

Approximately one-third of studies focussed on water-borne diseases (N=52, 32.9%) and vector-borne diseases (N=46, 29.1%), with roughly one quarter (N = 40, 25.3%) focussed on air-borne and sexually transmitted infections (STIs; Table 2). Ten studies were on blood-borne diseases (all Ebola Virus Disease (EVD) in DRC and Uganda) [64–66,72,78,107] and just three on vehicle-borne diseases (tetanus, norovirus, and intestinal parasites) [30,58,71].

**Table 2.** Key descriptive characteristics of studies (from 85 published research articles) included for data charting (N = 158). **N.B**. The greater number of studies than articles is due to the possibility of multiple studies and interventions being reported from individual articles

|  |  |
| --- | --- |
| ***Study design*** | ***n (%)*** |
| Case study | 57 (36.1%) |
| Cross-sectional | 38 (24.1%) |
| Pre-post | 38 (24.1%) |
| Controlled intervention | 17 (10.8%) |
| Case-control | 5 (3.2%) |
| Cohort | 3 (1.9%) |
| ***Disease transmission type*** |  |
| Water-borne | 52 (32.9%) |
| Vector-borne | 46 (29.1%) |
| Air-borne | 24 (15.2%) |
| Sexually-transmitted | 16 (10.1%) |
| Blood-borne | 10 (6.3%) |
| Multiple | 7 (4.4%) |
| Vehicle-borne | 2 (1.3%) |
| Not specified | 1 (0.6%) |
| ***NPI type*** |  |
| Community | 55 (34.8%) |
| Disease control | 44 (27.8%) |
| Personal | 38 (24.1%) |
| Environmental | 21 (13.3%) |
| ***Measure*** |  |
| Effectiveness | 81 (51.3%) |
| Feasibility | 53 (33.5%) |
| Acceptability | 24 (15.2%) |
| ***Research type*** |  |
| Quantitative | 94 (59.5%) |
| Qualitative | 64 (40.5%) |
| **Quality rating** |  |
| Good | 29 (18.4%) |
| Fair | 38 (24.1%) |
| Poor | 91 (57.6%) |
| **Total** | **158 (100%)** |

Approximately one-third of the studies (N = 55, 34.8%) were of community measures, most of which (N = 32) were studies of risk communication and community engagement (RCCE), and the bulk of the remainder assessments of WASH activities (Figure 3). Just two studies examined physical distancing, both during an EVD outbreak in DRC [64,65]. One quarter of the studies examined each of disease control (N = 44, 27.8%) and personal protective (N = 38, 24.1%) NPIs. Among the former, most studies (N = 33) investigated active case finding (ACF), while among the latter, half (N = 19) focused on vector protection and a quarter (N = 9) on hand hygiene. Of the 21 studies that explored environmental NPIs, the vast majority explored vector control, with a small number looking at cleaning, disinfection, and waste management.

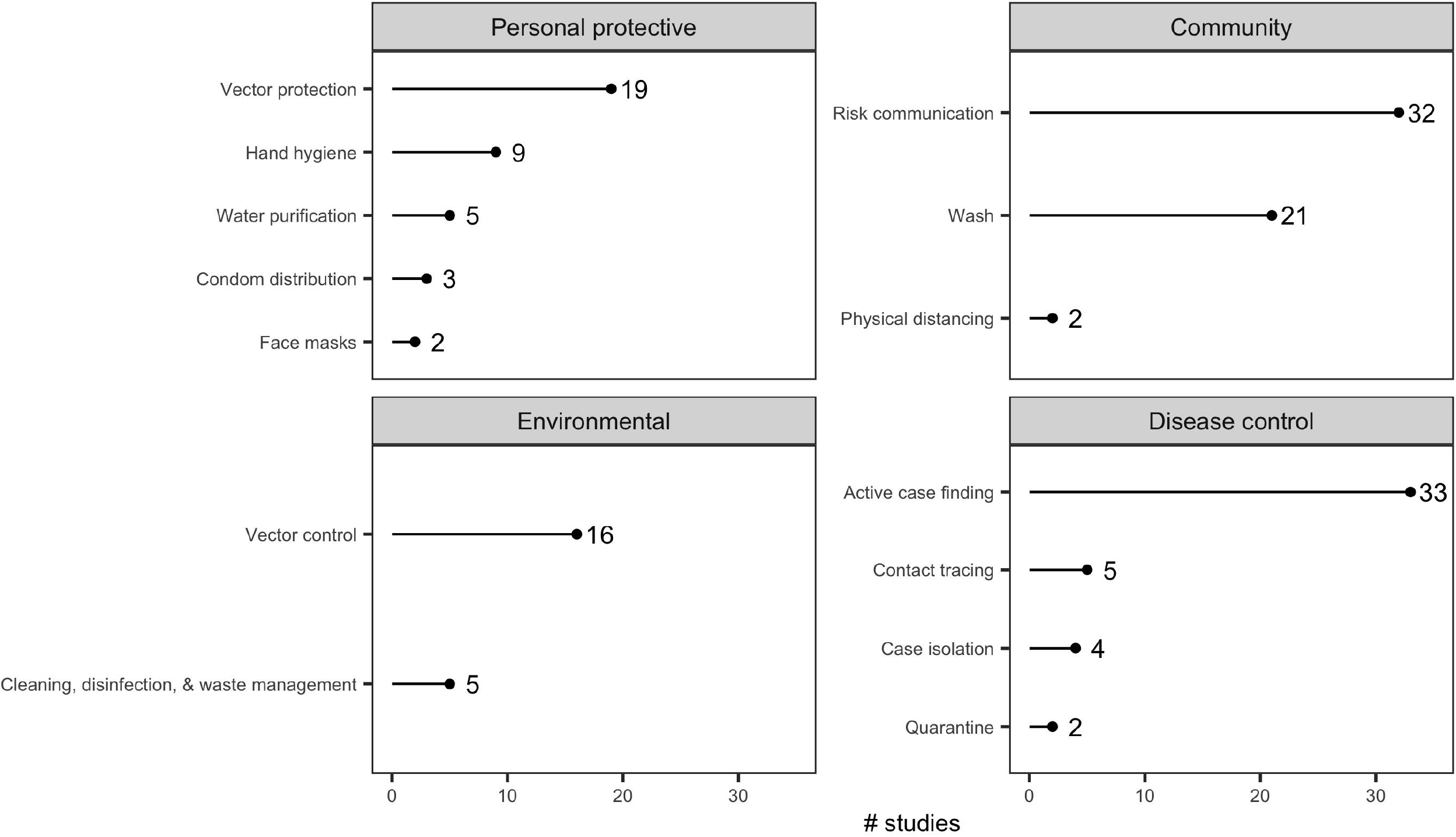

Assessments of effectiveness accounted for approximately half of the studies (N = 81, 51.3%), three-quarters of which were quantitative and one-quarter qualitative. Feasibility assessments accounted for one third of the studies, while the smallest category of research type was acceptability. Acceptability studies were mostly quantitative (N = 14, 58.3%), while feasibility studies were mostly qualitative (N = 32, 60.4%).

### Critical appraisal within sources of evidence

Case studies, the lowest ranking design on the quality rating scale, were the predominant study design (N = 57, 36.1%, Table 2). Cross-sectional studies and “pre-post evaluations” (studies with an ecological design comparing measures of interest before and after the introduction of an intervention but lacking a control group), each made up nearly one quarter of studies (N = 38, 24.1%), The highest-ranked study designs, namely controlled intervention, cohort, and case-control studies, accounted for just one-sixth of included studies (N = 25, 15.8%).

Most feasibility assessments used a case study design (N = 31, 58.5%), while most acceptability assessments were done by cross-sectional survey (N = 13, 54.2%), predominantly household survey or key informant interview (Supplementary Figure 1). Effectiveness studies had the greatest variation in study design, with sizable proportions of controlled intervention, pre-post, cross-sectional and case study designs.

Over half the studies were rated “poor” (N=91, 57.6%), with just one-fifth (N=29, 18.4%) rating “good” (Table 2). Approximately half of both the acceptability (N=12, 50%) and effectiveness (N=38, 46.9%) studies were rated as “poor”, with the remainder evenly split between “good” and “fair” ratings (Table S1, Supplementary Materials). Feasibility studies fared worse, with over three-quarters (N=41, 77.4%) rated as “poor” and just 4 (7.5%) achieving a “good” rating, likely due to the reliance on lower-quality case study designs to assess this measure.

Most interventions described were of short (under one year) duration, with a maximum of 12 years (IQR: 0-1, range: 0-12). The median delay between the start of the intervention and publication was three years (but with a very long-tailed distribution of up to 22 years) [IQR: 2-4, range: 0-22].

### Synthesis of results

#### Environmental measures

##### Vector control

Studies indicate indoor residual spraying (IRS) is a feasible intervention in complex emergencies [82], refugee camps [34,54,88] and post-disaster settings [33,96]. Important barriers included logistical problems such as absence of roads, remoteness of houses, and insecurity [82,96]. High coverage has been achieved in refugee settlements, where structured and close settlements facilitate IRS implementation [34].

While few studies assessed the acceptability of IRS, a cross-sectional survey within Afghan refugee settlements in Pakistan reported that incomplete coverage was due to the absence of household decision-makers during implementation [34].

Substantial evidence indicates IRS can (cost-)effectively reduce malaria incidence and mortality in camp and camp-like settings, including in Sudan [39], Pakistan [34,54,86,88] and Myanmar [114]. To ensure effectiveness, insecticide resistance, the timing of IRS implementation and malaria transmission, and the endophily and endophagy of the target vector must be considered [82,88]. Case-studies suggested that the application of insecticide (IRS, larvicide or fogging) can reduce exposure to a range of mosquito species following extreme weather events [33,96], though the impact is likely to be location-, intervention-, or vector species-specific [50].

##### Cleaning, disinfection & waste management

Quantitative evidence of feasibility of cleaning, disinfection and waste management was limited to a single case study of drinking water container disinfection during a shigellosis outbreak in an IDP camp in Sudan, with 88% of containers disinfected within five days [108]. Logistical issues, including stock shortages, have been qualitatively identified as a limiting factor in WASH intervention feasibility within IDP camps [74].

A cross-sectional study conducted during a cholera outbreak in post-earthquake Haiti found high acceptability of household disinfection kits (97.6% uptake) [44]. Qualitative evidence within multiple IDP camps and across various socio-cultural settings indicated concerns regarding the use of chlorine, including fears of poisoning or sterilisation and unfavourable smell which, when combined with ineffective communication, may reduce the acceptability of this NPI [74,108].

Evidence on the effectiveness of cleaning, disinfection and waste management on infectious disease transmission is highlighted by a case-control study in Dadaab refugee camp, which found that inadequate WASH interventions were associated with increased cholera risk [45].

#### Community measures

##### Risk communication

RCCE interventions were found to be highly feasible for communicating risk of cholera [83], hepatitis E [104], malaria [109], and HIV [102]. Sex and age differences were reported, with seeking treatment at health facilities, notifying sexual partners about symptoms, and adopting protective barriers well adopted among men but less among women, highlighting the importance of population-targeted messaging [40].

Effectiveness of RCCE was contextually dependent. Educational campaigns were generally highly effective for STIs. Community sensitisation programmes were associated with increased rates of HIV testing and case finding [76,83], while education and radio messaging campaigns improved STI risk awareness and behaviour [40,102,109]. However, STI education campaigns in Ngara refugee camp, Tanzania, were found to be ineffective, with no apparent impact on sexual behaviour and with STI prevalence increasing over time [68].

WASH RCCE campaigns were largely effective at improving hand-washing behaviour [73], knowledge of prevention strategies [94], and reductions in consumption of unsafe water [46] and water-borne illness morbidity and mortality [46,59,70,81,103]. Waste disposal education was also associated with reduced latrine blockages in an informal settlement in Dhaka [112]. However, awareness campaigns did not lead to a change in water purification behaviours in Kathmandu [95].

RCCE campaigns, in conjunction with insecticide-treated net (ITN) distribution, were highly effective at improving knowledge on the causes of malaria and correct use of ITNs [84,99], and education campaigns aimed at improving awareness of dengue led to an increase in knowledge of the disease and vector [31]. Education campaigns for tetanus were found to substantially increase vaccine uptake in a refugee camp in Darfur, Sudan [30]. RCCE was implemented with success for tuberculosis (TB) treatment, reducing default [98] and increasing health seeking behaviour [93].

During an EVD outbreak, a majority of respondents to a qualitative survey believed that RCCE contributed to the response efforts [64], although one study in the same setting reported difficulties in communicating health information due to language barriers, which, once addressed, improved RCCE effectiveness [72]. Language barriers were also reported to have led to community mistrust and decreased effectiveness of cholera outbreak interventions [74].

##### Water, sanitation, and hygiene (WASH)

Establishing WASH services was often feasible, but financial constraints [32], sustainable maintenance [113], and implementation were important challenges reported [45].

Establishing trust between stakeholders was identified as a key determinant of successful implementation [35].

One study explored the acceptability of WASH interventions in six IDP camps in Borno State, Nigeria [74]. Overcrowding and distance from home caused lack of acceptability of communal latrines, while chlorine use required the mobilisation of trusted members of the community to enhance acceptability.

A programme that complemented WASH interventions with hygiene education led to improvements in household water quality, access to sanitation, and improved hand washing behaviour, associated with reduced diarrhoea incidence among children [73]. WASH interventions were associated with low cholera outbreak case fatality ratio [74]. In a case study post-Tsunami in Indonesia, no major outbreaks of any disease occurred, possibly linked to hygiene interventions [70].

##### Physical distancing

We identified just two studies exploring physical distancing as an NPI, both of which were cross-sectional studies of the acceptability of these measures during an EVD outbreak response in DRC. They reported low acceptability of burial practices that excluded touching the corpse [64,65] and a willingness to hide suspected Ebola-positive family members from health authorities [64].

#### Personal protective measures

##### Vector protection

Personal vector protection measures (ITN and mosquito repellent) were reported to be both highly feasible [96] and acceptable [53], with user costs being the only important barrier noted. ITNs were highly effective in reducing malaria infection risk in various settings [87,89,91,114].

##### Hand hygiene

The feasibility of implementing hand hygiene measures was context-specific, with uptake of measures and a reduction in disease incidence seen in shelters following natural disasters [51] and among refugees displaced by conflict [104]. Conversely, substantial issues with feasibility were observed during a norovirus outbreak in a refugee camp in Germany, owing to a lack of water supply, alcohol-based sanitizer, and language barriers [48]. Acceptability issues due to cultural differences in eating habits were also reported [48]. Hand hygiene, in conjunction with other preventative measures, was found to be effective at reducing the influenza A attack rate during an outbreak in a temporary shelter following the Tōhoku earthquake in 2011 [57], and no infectious disease outbreaks were detected following the 2004 tsunami in Aceh, Indonesia where soap and hygiene kits were distributed [70]. Age-differentiated effectiveness was also reported, with handwashing effective against dysentery among children aged over 5, but not those younger, in an informal settlement in Kolkata [97].

##### Water purification

Bottled water distribution in a temporary shelter following an earthquake in Sichuan Province, China was feasible in the short term, but sustainability was problematic [113], while water purification and the distribution of improved water containers was highly effective at reducing the occurrence of diarrhoea among refugees in Africa [42,46,85].

##### Face masks

Face mask distribution was found to be feasible, acceptable, and effective when used in conjunction with other control measures aimed at preventing and reducing the spread of Influenza A among populations living in temporary shelters following natural disasters in Japan [51,57].

##### Condom distribution

Condom distribution had mixed effectiveness, with one study finding a substantial reduction in high-risk sexual behaviours among refugees in Liberia and Sierra Leone [109], while another found no effect on sexual behaviour among Rwandan refugees, with an increase in STIs reported over the intervention period [68].

#### Disease control

##### Active case finding

Active case finding (ACF) for HIV and TB was judged to be both feasible [75,76,106] and acceptable [76] in refugee camps in Sudan and Uganda, while ACF of diarrhoeal disease by household visitation and mobile phone surveillance was feasible, but limited by technical issues, lack of treatment follow-up and by poor information sharing and coordination [32,38]. For case detection of human African trypanosomiasis, passive screening at health facilities was easier to implement than ACF among refugees in South Sudan [79], while widespread ACF among a rural, non-displaced conflict-affected population in the DRC was prevented by conflict and instability [105].

Desire for knowledge of HIV status, peer encouragement, and provision of a “package” (including RCCE and bundling with tests for other diseases) were key to the high acceptability of HIV voluntary counselling and testing (VCT) observed in informal settlements [56,83]. ‘Opt-out’ and community-based screening strategies increased access and uptake, particularly among high-risk groups [61,76,83]. In a refugee camp in Sudan, refusal of TB screening was associated with a fear of negative consequences of a positive test result, linked to the status of refugees within the host country and lack of trust [106].

ACF contributed to improved TB detection, defaulting rates, and treatment outcomes among TB patients in informal settlements and among refugees [41,67,77]. In IDP camps in the Central African Republic, screening for malaria by rapid diagnostic test (RDT) led to a very high percentage (98%) of positive cases receiving appropriate treatment [92]. In a refugee camp in Tanzania, a combined ACF and RCCE approach ensured that most cholera patients arrived in a stable condition, improving their chances of survival [81]. Similarly, a mixed ACF and RCCE programme resulted in more refugees screened and more HIV+ patients detected in a refugee camp in Uganda [75]. Pre-departure screening of refugees from the Thai-Burmese border resulted in a substantial decrease in helminth infections and moderate-to-severe anaemia [71].

##### Contact tracing

Contact tracing was feasible for the detection and management of an influenza A outbreak in a post-disaster setting in the USA [63]. It was also feasible and effective for TB control among Syrian refugees in Jordan [52] and in an informal settlement in Haiti [67], leading to the identification of additional cases of latent TB.

##### Case isolation

In conflict-affected settings in Central Africa, isolation of EVD cases was reported to have low acceptability [65], despite reducing mortality and leading to shorter outbreak duration [78]. Case isolation, in combination with other measures including personal hygiene and mask wearing, was found to be effective at reducing the attack rate of influenza A during an outbreak in temporary shelters in Japan [57].

##### Quarantine

The sole study on quarantining reported that a community-imposed quarantine on household members of an uncooperative EVD case successfully prevented the spread of the disease [78]. Further details were unavailable.

## Discussion

### Summary of evidence

The evidence-base for the feasibility, acceptability, and effectiveness of NPIs in crisis-affected populations and informal settlements was limited for all but a small number of communicable diseases. Most studies were of short duration (typically under one year) and produced low-quality evidence due to study design limitations. While this is understandable given the complexity and insecurity of humanitarian emergencies, it remains an important shortcoming of the body of evidence, limiting what can be asserted and the generalisability of the findings, and underscoring the need for more and better-quality research in different contexts.

The research was limited to a narrow range of settings, transmission modes, NPIs, and timespan. Most research was conducted in those parts of sub-Saharan Africa and Asia in which most crisis-affected people live, and the high number of studies in conflict- and disaster-settings reflects the predominance of these crises. Furthermore, as has been described elsewhere [20], approximately half of the studies were in camp or camp-like settings, despite most refugees and IDPs living in out-of-camp settings. There was scant evidence from informal settlements (slums), which is concerning given the large number of affected people and lack of historical health data [17,18].

The preponderance of research on water-borne and vector-borne diseases may reflect the types of interventions available and typically implemented by humanitarian relief agencies (e.g., WASH, ITNs) and the relative ease of implementing and studying them. However, air-borne diseases remain vastly under-investigated, particularly given their potential for large, highly transmissible epidemics of devastating impact. Such modes of transmission may necessitate novel and difficult-to-implement approaches such as quarantines, travel restrictions, and restrictions of (mass) gatherings of which the feasibility, acceptability, and ultimately the effectiveness are currently poorly documented in crisis-affected populations and informal settlements.

Despite the limited body of evidence, and the large extent to which the evidence was dependent on the context in which the NPIs were implemented, it was possible to identify some common themes that emerged from the scientific literature. Relatively simple and cost-effective interventions, such as community-level environmental cleaning and provision of WASH services, and those that involved the distribution of items for personal protection such as ITNs, were both highly feasible and acceptable, which aided in their effectiveness. The principal barriers with such interventions tended to be logistical, financial, or human resource constraints that impacted access and feasibility, particularly over the long term beyond the acute phase of a crisis. Conversely, resource-intensive individual-level interventions that involve potential restrictions on personal liberty, such as active finding and isolation of cases and contact tracing, were more challenging to implement, and community trust played an extremely important role in their acceptance and success.

Vertical programmes imposed on communities by external actors have a mixed history of success, and many studies identified low acceptability and poor feasibility in NPI maintenance over the medium- and long-term. An alternative model is increasingly being sought, with humanitarian actors working more closely with communities to implement sustainable programmes with viability over the long-term and complementing development programmes [115]. To support this view, the evidence suggested that many NPIs benefited from incorporating an RCCE component. Regular and meaningful contact with affected communities facilitated feedback loops, which may have helped to identify potential concerns and propose community-led solutions [116]. Moreover, NPIs should be considered in the wider context in which they are implemented, which has the potential to impact all three measures of success. For example, interventions that restrict people’s ability to carry on their regular activities, including work and attending social gatherings, can have severe consequences for their mental and physical well-being. Thorough consideration should be given to mitigate such impacts, such as through providing mental health and psychosocial support and home-care packages that include essential food and non-food items [116].

### Limitations

We only included articles published in peer-reviewed journals, but the nature of conducting research in crisis settings, much of which is done as operational research to inform programmes, may result in a substantial wealth of unpublished “grey” literature. The English language restriction further limited the pool of available studies.

The findings are likely to be subject to publication bias, with interventions that were found to be less feasible or effective less likely to be published. It cannot be stressed enough that negative findings should also be published, to avoid “reinventing the wheel”, with subsequent researchers repeating work that was inadequately documented.

Therefore, we encourage researchers with unpublished data to publish their research, and for their employers to provide the time necessary to complete this work, so that important evidence is documented and made widely accessible.

## Conclusions

The implementation of NPIs poses unique practical constraints in resource-limited settings that need to be considered when planning disease prevention and control programmes. The evidence-base in the peer-reviewed literature on this important theme is scant, occasionally contradictory, and generally low quality. The evidence found through this review revealed major limitations, and cautious interpretation is therefore needed, particularly where the quality of evidence is weakest.

Implementing rigorous, high quality studies in these complex and insecure environments is both logistically and ethically challenging [117–119]. However, it is important that guidance on infectious disease prevention and control is rooted in a firm evidence-base, i.e., with high-quality studies. More thought should also be given to how useful lessons can be gathered from studies among stable populations with similar profiles, such as informally housed, non-displaced crisis-affected populations, and refugees and IDPs in protracted crisis settings. Given the substantial resources engaged in protecting populations in emergency situations, basing these on better quality evidence would be a valuable use of resources and should be increasingly considered.

New evidence is beginning to emerge from implementing NPIs in the context of COVID-19, including in crisis-affected populations and informal settings [12,13,120–125], which is encouraging. We call for similar work to be conducted and documented to help us better prepare for current and future infectious diseases outbreaks in these contexts.

## Supporting information

Full bibliographic search terms

## Data Availability

All data generated or analysed during this study are included in this published article (and its supplementary information files).

## Acknowledgments

SB, KF, AH, JS and IJS acknowledge funding from the MRC Centre for Global Infectious Disease 514 Analysis (reference MR/R015600/1), jointly funded by the UK Medical Research Council 515 (MRC) and the UK Foreign, Commonwealth & Development Office (FCDO), under the 516 MRC/FCDO Concordat agreement and is also part of the EDCTP2 programme supported 517 by the European Union. SB acknowledges funding from the Wellcome Trust (219415). JS acknowledges funding from the Wellcome Trust (grant reference: 215163/Z/18/Z). IJS was supported by the QMEE CDT, funded by NERC grant number NE/P012345/1.

The authors alone are responsible for the views expressed in this article and they do not necessarily represent the views, decisions, or policies of the institutions with which they are affiliated.

