## Supplementary material for "Feasibility, Acceptability, and Effectiveness of Non-Pharmaceutical Interventions for Infectious Disease Prevention and Control in Crisis-Affected Settings and Informal Settlements: A Scoping Review": Table S1

Table S1: Article inclusion and exclusion criteria for scoping review

| Category | Included | Excluded |
| --- | --- | --- |
| Population of interest | All individuals living in humanitarian settings and/or informal housing (slums). This includes refugees, internally displaced persons, asylum seekers, the informally housed, prisoners of war, and non-displaced (but crisis-affected) persons | Populations living in fragile settings. This includes immigrants, indigenous people, neglected high-risk groups including IV drug users and sex workers, and prisoners and other detainees (except prisoners of war) |
| Intervention | NPIs against infectious diseases | No NPIs against infectious diseases mentioned |
| Article type | Any quantitative or qualitative study describing an NPI and which contributes primary data. Acceptable article types were primary research, narrative reports, and case studies | Any study with no specific NPI or that only describes needs, prevalence, risk factors, or results from modelling secondary data. Excluded article types were editorials and opinion pieces, modelling studies, and review articles (which were screened for references but were themselves excluded from the review) |
| Crisis type | Any acute or protracted armed conflict, natural disaster, complex emergency, and informal settlement (slum) | Studies conducted before a crisis has occurred (pre-crisis) or during the post-emergency (recovery) phase, in fragile settings, or in epidemics in otherwise non-crisis settings |
| Setting | Refugee camps, Conflict-affected, Natural disaster, Informal settlement (slum), Hosted (refugees/IDPs) | Prisons, detention centres (including asylum centres), migrant screening (unless screening within a crisis-affected host country) |
| Publication date | 1970–2020 | Research published prior to 1970 or after 2020 |
| Language | English | Other languages |
