## Supplementary material for "Feasibility, Acceptability, and Effectiveness of Non-Pharmaceutical Interventions for Infectious Disease Prevention and Control in Crisis-Affected Settings and Informal Settlements: A Scoping Review": Full bibliographic search terms

(("non-pharmaceutical") OR ("nonpharmaceutical") OR ("intervention") OR ("social distancing") OR ("school closures") OR ("workplace closures") OR ("mass gatherings") OR ("hygiene") OR ("spraying") OR ("bednet") OR ("bed net") OR ("bed-net") OR ("vector-control") OR ("vector control") OR ("WASH") OR ("contact tracing") OR ("travel restriction") OR ("quarantine") OR ("public health campaigns") OR ("public health and social measures") OR ("screening") OR ("curfew") OR ("face mask") OR ("physical distancing")).

AND (("infectious") OR ("epidemic") OR ("pandemic") OR ("outbreak") OR ("transmission")),

AND (("refugee") OR ("humanitarian") OR ("Internally displaced person") OR ("Internally displaced people") OR ("camp") OR ("slum") OR ("migration") OR ("disaster") OR ("war") OR ("prison") OR ("jail") OR ("armed conflict") OR ("detention centre") OR ("crisis")).
