## Supplementary figures and images for "Feasibility, Acceptability, and Effectiveness of Non-Pharmaceutical Interventions for Infectious Disease Prevention and Control in Crisis-Affected Settings and Informal Settlements: A Scoping Review"

### Figure S1

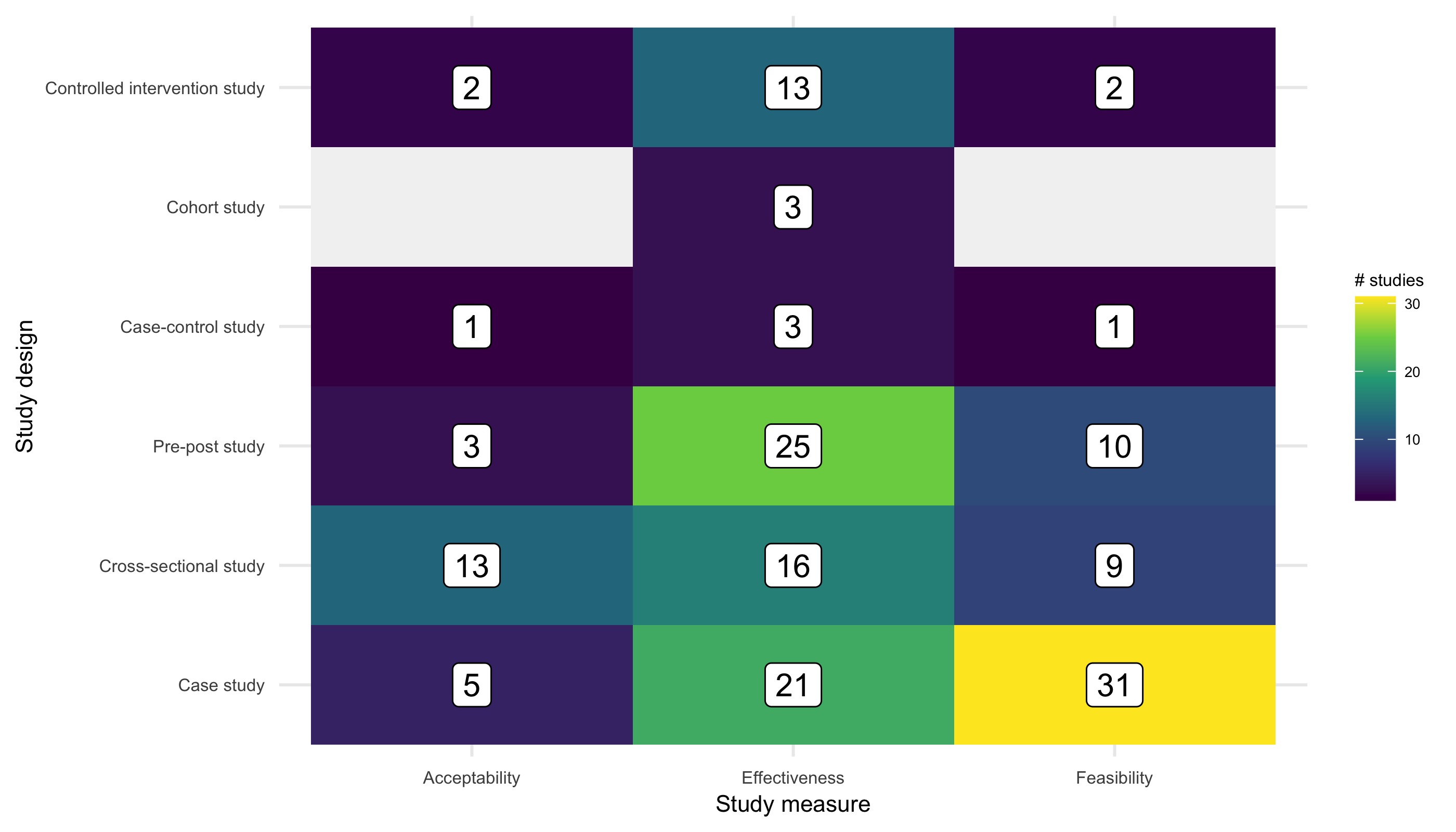
