## Supplementary material for "Feasibility, Acceptability, and Effectiveness of Non-Pharmaceutical Interventions for Infectious Disease Prevention and Control in Crisis-Affected Settings and Informal Settlements: A Scoping Review": Table S2

Table S2: Summary information from 158 research studies (from 85 published research articles) included for data charting.

| **Reference** | **Country** | **Crisis type** | **Population** | **Shelter** | **Transmission mode** | **Disease** | **Level of intervention** | **NPI** | **Intervention** | **Study design** | **Measure** | **Research type** | **Study size** | **Quality** | **Key findings** |
| --- | --- | --- | --- | --- | --- | --- | --- | --- | --- | --- | --- | --- | --- | --- | --- |
| Adam et al., 2015 | Sudan | conflict | internally displaced | camp or camp-like | vehicle-borne | tetanus | community | risk communication | Mass education campaigns on tetanus toxoid vaccination services | pre-post study | effectiveness | quantitative | 88,984 | good | Education campaign was effective at increasing tetanus toxoid vaccination awareness and uptake |
| AhbiRami & Zuharah, 2020 | Malaysia | natural disaster | non-displacement | non-displacement | vector-borne | dengue | community | risk communication | KAP study on the effectiveness of dengue health education program | pre-post study | effectiveness | quantitative | 203 | good | Education campaign was effective at increasing knowledge |
| Altmann et al., 2017 | Yemen | conflict | non-displacement | non-displacement | water-borne | cholera | community | risk communication | Training of CHWs & hygiene promoters to provide hygiene education | case study | feasibility | qualitative | 27801 | poor | Risk communication feasible at both water distribution points and in the household. |
| Altmann et al., 2017 | Yemen | conflict | non-displacement | non-displacement | water-borne | cholera | community | WASH | Provision of water to communities by trucking. | case study | feasibility | qualitative | 2430 HHs | poor | Water distribution feasible to large number of people, but safe storage was identified as potential concern. Financial constraints limited widespread feasibility |
| Altmann et al., 2017 | Yemen | conflict | non-displacement | non-displacement | water-borne | cholera | disease control | active case finding | Active case finding within the community. | case study | feasibility | qualitative | 7598 HHs | poor | Active case finding was feasible, but limited by poor information sharing and coordination. |
| Aumentado et al., 2015 | Philippines | natural disaster | not specified | not specified | vector-borne | dengue | environmental | vector control | Vector control measures (fogging, larviciding, search-and-destroy) | pre-post study | effectiveness | quantitative | ~2200 | poor | Reductions in vector densities observed in most villages |
| Aumentado et al., 2015 | Philippines | natural disaster | not specified | not specified | vector-borne | dengue | environmental | vector control | Vector control measures (fogging, larviciding, search-and-destroy) | case study | feasibility | qualitative | ~2200 | poor | Vector control operations hindered by lack of HR, logistical support, and community participation in assessments. |
| Bouma et al., 1996 | Pakistan | conflict | refugee | camp or camp-like | vector-borne | malaria | environmental | vector control | Indoor residual spraying of tents with insecticide | pre-post study | acceptability | quantitative | 26,119 | good | IRS was acceptable |
| Bouma et al., 1996 | Pakistan | conflict | refugee | camp or camp-like | vector-borne | malaria | environmental | vector control | Indoor residual spraying of tents with insecticide | pre-post study | effectiveness | quantitative | 26,119 | good | P. falciparum prevalence reduced following IRS |
| Brocklehurst et al., 2013 | Zimbabwe | complex emergency | non-displacement | non-displacement | water-borne | cholera | community | WASH | Supply of water purification products, unblocking sewage pipes, training staff, improving sanitation infrastucture and risk communication | case study | feasibility | quantitative | Not stated | poor | WASH campaign reached over 30% of Zimbabwe's population |
| Brocklehurst et al., 2013 | Zimbabwe | complex emergency | non-displacement | non-displacement | water-borne | cholera | community | WASH | Supply of water purification products, unblocking sewage pipes, training staff, improving sanitation infrastucture and risk communication | case study | feasibility | qualitative | Not stated | poor | Significant trust between stakeholders identified as key to success |
| Brooks et al., 2017 | Democratic Republic of the Congo | conflict | internally displaced | camp or camp-like | vector-borne | malaria | personal protective measures | vector protection | Universal free bednet distribution for malaria prevention | cross-sectional survey | feasibility | qualitative | 55 | poor | High community awareness of and access to bednets, but living conditions reduced feasibility of use. |
| Brooks et al., 2017 | Democratic Republic of the Congo | conflict | internally displaced | camp or camp-like | vector-borne | malaria | personal protective measures | vector protection | Universal free bednet distribution for malaria prevention | cross-sectional survey | feasibility | quantitative | 411 | poor | Despite free ITN provision, usage was low (~20%). |
| Burns et al., 2012 | Sierra Leone | conflict | refugee | camp or camp-like | vector-borne | malaria | personal protective measures | vector protection | Provision of insecticide-treated plastic sheeting (ITPS) for shelter | cohort study | effectiveness | quantitative | 1610 | fair | Full ITPS coverage was effective at deterring parasites, while only having ITPS on the roof was not effective. Full ITPS shelter coverage and roof-only ITPS coverage both reduced malaria incidence, but full ITPS shelter coverage was more effective. |
| Carstensen et al., 2019 | Bangladesh | informal housing | informally housed | informal housing | water-borne | diarrhoeal | disease control | active case finding | Diarrhoea surveillance via mobile phone | cross-sectional survey | feasibility | qualitative | 476 HHs | poor | Mobile phone surveillance of diarrhoeal disease was feasible. Lack of treatment follow-up and technical issues reduced incentive to report. |
| Charlwood et al., 2001 | Sudan | conflict | refugee | camp or camp-like | vector-borne | malaria | environmental | vector control | IRS (malathion) of walls | controlled intervention study | effectiveness | quantitative | 48715 | poor | IRS reduced malaria mortality among under 5s |
| Chen et al., 2008 | Guinea | conflict | refugee | camp or camp-like | sexually-transmitted | not specified | community | risk communication | Health education about STI prevention | cross-sectional study | effectiveness | quantitative | 889 | good | Health education was effective in improving STI knowledge and behaviour. Seeking treatment at health facilities, notifying sexual partners about symptoms and adopting protective barriers were well adopted among men but less among women |
| Chen et al., 2008 | Guinea | conflict | refugee | camp or camp-like | sexually-transmitted | not specified | personal protective measures | condom distribution | Provision of contraception | cross-sectional study | effectiveness | quantitative | 889 | good | Condom use was infrequently reported. |
| Cookson et al., 2015 | Jordan | conflict | refugee | camp or camp-like; hosted | air-borne | tuberculosis | disease control | active case finding | Screening for TB and TB awareness sessions | case study | effectiveness | quantitative | 60000 | fair | ACF enhanced TB detection (40% increase in detection) |
| Doocy & Burnham, 2006 | Liberia | conflict | internally displaced | camp or camp-like | water-borne | diarrhoeal | personal protective measures | water purification | Improved water sources and disinfectant for water purification | controlled intervention study | effectiveness | quantitative | 400 HHs | fair | Disinfectant significantly reduced diarrhoea |
| Elledge et al., 2007 | United States of America | natural disaster | internally displaced | camp or camp-like | water-borne | gastroenteritis | community | WASH | Provision of various WASH interventions. Staff, volunteers, and evacuees educated on handwashing. Provision of gel hand sanitizers. | case-study | feasibility | qualitative | 1 evacuation centre | poor | WASH facilities were insufficient and overwhelmed. Lack of hygienic practices observed. |
| Gartley et al., 2013 | Haiti | natural disaster | non-displacement | non-displacement | water-borne | cholera | environmental | cleaning, disinfection, & waste management | Household disinfection kits distributed to patients in CTC. | cross-sectional survey | acceptability | quantitative | 208 | poor | Disinfection kits were acceptable and used by recipients |
| Golicha et al., 2018 | Kenya | conflict | refugee | camp or camp-like | water-borne | cholera | community | WASH | Provision of latrines, handwashing facilities, chlorine and soap. Risk communication. | case-study | feasibility | quantitative | ~300,000 | poor | Infeasible to implement WASH to required standards |
| Golicha et al., 2018 | Kenya | conflict | refugee | camp or camp-like | water-borne | cholera | environmental | cleaning, disinfection, & waste management | Provision of latrines, handwashing facilities, chlorine and soap. Risk communication. | case-control study | effectiveness | quantitative | 96 | fair | Soap use & owning a household latrine led to non-significant reduction in cholera OR. Shared and communal latrines were more risky than open defecation. |
| Graf et al., 2010 | Cameroon | informal housing | informally housed | informal housing | water-borne | diarrhoeal | community | risk communication | Education campaign | controlled intervention study | effectiveness | quantitative | 2976 HHs | fair | Solar-powered water purification and education campaign reduced diarrahoea and consumption of unsafe water |
| Graf et al., 2010 | Cameroon | informal housing | informally housed | informal housing | water-borne | diarrhoeal | personal protective measures | water purification | Distribution of solar-powered water purification devices | controlled intervention study | effectiveness | quantitative | 2976 HHs | fair | Solar-powered water purification and education campaign reduced diarrahoea and consumption of unsafe water |
| Graham et al., 2002 | Pakistan | conflict | refugee | camp or camp-like | vector-borne | malaria | personal protective measures | vector protection | Treatment of bedsheets with insecticides | controlled intervention study | acceptability | quantitative | 88 HHs | poor | High acceptability of insecticide-treated sheets. Deltamethrin less acceptible than other insecticides due to skin irritation. |
| Grote et al., 2017 | Germany | conflict | refugee | camp or camp-like | water-borne | norovirus | personal protective measures | hand hygiene | Provision of hand sanitisers and hand hygiene promotion. Sanitary facility cleaning increased and staff educated on hygiene measures. | case study | acceptability | qualitative | 982 | poor | Differences in eating habits and hygiene standards resulted in variable acceptability and uptake. |
| Grote et al., 2017 | Germany | conflict | refugee | camp or camp-like | water-borne | norovirus | personal protective measures | hand hygiene | Provision of hand sanitisers and hand hygiene promotion. Sanitary facility cleaning increased and staff educated on hygiene measures. | case study | feasibility | qualitative | 982 | poor | Implementation was challenging because of lack of water supply, ineffective alcohol-based hand sanitizers needed to be replaced, hygiene risk communication needed to be delivered in many languages, and general avoidance of shared facilities was infeasible. |
| Hamze et al., 2016 | Democratic Republic of the Congo | conflict | internally displaced | camp or camp-like | vector-borne | malaria | disease control | active case finding | Community screening conducted by CHWs | case-control study | effectiveness | quantitative | ~40,000 | fair | Screening household contacts of malaria cases was not an efficient case-finding strategy. Symptom-based screening may be a simpler and cost-effective method to identify individuals at increased risk of malaria. |
| Harris et al., 2014 | United States of America | natural disaster | non-displacement | non-displacement | vector-borne | NA | environmental | vector control | Aerial insecticide spraying | pre-post study | effectiveness | quantitative | 8 sites | poor | Mosquitoes abundance dropped (non-significantly) following spraying. |
| Hatta et al., 2012 | Japan | natural disaster | internally displaced | camp or camp-like | air-borne | influenza a | disease control | case isolation | Isolation of symptomatic patients until 2 days post-fever | case study | feasibility | qualitative | 1360 | poor | Feasible to isolate cases, but not quantified |
| Hatta et al., 2012 | Japan | natural disaster | internally displaced | camp or camp-like | air-borne | influenza a | personal protective measures | face masks | Distribution of facemasks | case study | feasibility | qualitative | 1360 | poor | Feasible to distribute facemasks, but not quantified |
| Hatta et al., 2012 | Japan | natural disaster | internally displaced | camp or camp-like | air-borne | influenza a | personal protective measures | hand hygiene | Distribution of hand sanitiser | case study | feasibility | qualitative | 1360 | poor | Feasible to install alcohol-based hand sanitiser, but lack of running water was obstacle |
| Hosten et al., 2018 | Jordan | conflict | refugee | not specified | air-borne | tuberculosis | disease control | contact tracing | Contact tracing for screening | cross-sectional survey | effectiveness | quantitative | 481 | fair | Contact tracing led to identification of additional cases of latent TB |
| Hosten et al., 2018 | Jordan | conflict | refugee | not specified | air-borne | tuberculosis | disease control | contact tracing | Contact tracing for screening | cross-sectional survey | feasibility | quantitative | 481 | fair | Roughly half of contacts were clinically assessed within 120 days of diagnosis of primary case, of which ~90% were treated. Among those reached, intervention was highly feasible. |
| Howard et al., 2010 | Afghanistan | conflict | non-displacement | non-displacement | vector-borne | malaria | personal protective measures | vector protection | ITN usage assessed using FGDs, in-depth interviews, and quantitative household survey | cross-sectional survey | acceptability | qualitative | 414 HHs | poor | ITNs were highly acceptable. Primary barrier was cost. Responsibility for purchasing decisions rests with the household head (usually male), but preferential use given to women and children. |
| Howard et al., 2017 | Pakistan | conflict | refugee | camp or camp-like | vector-borne | malaria | environmental | vector control | Vector control by annual IRS | pre-post study | effectiveness | quantitative | 2400000 | good | IRS was (cost-)effective against malaria in this low-endemicity setting (more cost-effective when incidence was higher). |
| Husain et al., 2015 | Ethiopia | conflict | refugee | camp or camp-like | water-borne | diarrhoeal | personal protective measures | hand hygiene | Household distribution of "handwashing bag" (with pictorial instructions, spigot and soap) | pre-post study | acceptability | quantitative | 222 HHs | good | Bag was highly acceptable. |
| Husain et al., 2015 | Ethiopia | conflict | refugee | camp or camp-like | water-borne | diarrhoeal | personal protective measures | hand hygiene | Household distribution of "handwashing bag" (with pictorial instructions, spigot and soap) | pre-post study | feasibility | quantitative | 222 HHs | good | Bags were durable (68% functional after 3 months). |
| Kabiru et al., 2011 | Kenya | informal housing | informally housed | informal housing | sexually-transmitted | hiv/aids | disease control | active case finding | Active case finding for HIV | cross-sectional study | acceptability | qualitative | 4058 | good | Not being sexually active and perception of being at low risk were main reasons for not wanting test. Desire to know or not know status was important in decision making. Encouragement from others (peers/counsellors/family) played an important minor role. |
| Kamigaki et al., 2014 | Japan | natural disaster | internally displaced | camp or camp-like | air-borne | influenza a | disease control | case isolation | Isolation of cases within designated areas of evacuation centres. | case study | effectiveness | quantitative | 1810 | poor | The influenza A attack rate was low (ranging between 0.8% and 7.7% for the 5 evacuation centres), possibly due to a combination of NPIs. |
| Kamigaki et al., 2014 | Japan | natural disaster | non-displacement | camp or camp-like | air-borne | influenza a | personal protective measures | face masks | Distribution of masks and frequent encouragement of use. | case study | acceptability | quantitative | 1810 | poor | High acceptability and uptake of mask wearing |
| Kamigaki et al., 2014 | Japan | natural disaster | non-displacement | camp or camp-like | air-borne | influenza a | personal protective measures | hand hygiene | Distribution of hand washing (using alcohol-based hand sanitizers or running water) and frequent encouragement of use. | case study | acceptability | quantitative | 1810 | poor | High acceptability and uptake of hand washing |
| Karmarkar et al., 2020 | United States of America | natural disaster | internally displaced | camp or camp-like | water-borne; food-borne; vehicle-borne | norovirus | community | WASH | Improved environmental and kitchen practices; hand hygiene; facility cleanliness; self-service practices for food and beverages; cleanliness of child play areas. | pre-post study | feasibility | quantitative | 6 shelters | poor | Rapid scale-up of WASH activities was feasible |
| Karmarkar et al., 2020 | United States of America | natural disaster | internally displaced | camp or camp-like | water-borne; food-borne; vehicle-borne | norovirus | disease control | active case finding | Introduction and application of comprehensive screening protocols | pre-post study | feasibility | quantitative | 6 shelters | poor | Rapid scale-up of screening activities was feasible |
| Khan et al., 2018 | Nepal | natural disaster | multiple | multiple | water-borne | cholera | community | risk communication | WASH interventions and education campaigns | case study | effectiveness | qualitative | Not stated | poor | No cholera epidemic in the aftermath of the earthquake |
| Khan et al., 2018 | Nepal | natural disaster | multiple | multiple | water-borne | cholera | community | WASH | WASH interventions and education campaigns | case study | effectiveness | qualitative | Not stated | poor | No cholera epidemic in the aftermath of the earthquake |
| Kimani et al., 2006 | Kenya | conflict | refugee | camp or camp-like | vector-borne | malaria | personal protective measures | vector protection | Clothes and beddings of the participants in treatment group treated with insecticide every after three weeks during study period. | controlled intervention study | acceptability | quantitative | 198 | fair | Almost all (12/14) households accepted insecticide treating |
| Kimani et al., 2006 | Kenya | conflict | refugee | camp or camp-like | vector-borne | malaria | personal protective measures | vector protection | Clothes and beddings of the participants in treatment group treated with insecticide every after three weeks during study period. | controlled intervention study | effectiveness | qualitative | 198 | fair | Intervention led to reduced mosquito bites, bedbugs, and lice |
| Kimani et al., 2006 | Kenya | conflict | refugee | camp or camp-like | vector-borne | malaria | personal protective measures | vector protection | Clothes and beddings of the participants in treatment group treated with insecticide every after three weeks during study period. | controlled intervention study | effectiveness | quantitative | 198 | fair | Intervention was effective at reducing malaria incidence |
| Kohli et al., 2012 | Democratic Republic of the Congo | conflict | non-displacement | non-displacement | sexually-transmitted | hiv/aids | disease control | active case finding | HIV testing at mobile clinic for female patients | cross-sectional survey | acceptability | quantitative | 772 | poor | Testing was highly acceptable (93%) |
| Lantagne & Clasen, 2013 | Haiti | natural disaster | non-displacement | non-displacement | water-borne | cholera | personal protective measures | water purification | Household water treatment and safe storage (HWTS) products distributed to households. | cross-sectional survey | effectiveness | quantitative | 363 HHs | poor | DSI Aquatabs Safe Storage Program was found to be the most effective at reducing E. coli contamination in water for both acute and recovery phases |
| Lantagne & Clasen, 2013 | Haiti | natural disaster | non-displacement | non-displacement | water-borne | cholera | personal protective measures | water purification | Household water treatment and safe storage (HWTS) products distributed to households. | cross-sectional survey | feasibility | quantitative | 363 HHs | poor | Household distribution of water purifying material only reached <1% affected population, while tanker truck water distributions reached 37 times more people. Successful programs targeted households with contaminated water, provided an HWTS method that effectively treated the water, had a population that was familiar with the product and willing to use it along with training in its use with the necessary supplies, including a safe storage container. |
| Liu et al., 2019 | United States of America | natural disaster | internally displaced | camp or camp-like | air-borne | influenza a | community | risk communication | Risk communication | case study | feasibility | qualitative | 7409 | poor | Risk communication was feasible to implement. |
| Liu et al., 2019 | United States of America | natural disaster | internally displaced | camp or camp-like | air-borne | influenza a | disease control | active case finding | Active case finding | case study | feasibility | qualitative | 7409 | poor | ACF was feasible to implement. |
| Liu et al., 2019 | United States of America | natural disaster | internally displaced | camp or camp-like | air-borne | influenza a | disease control | quarantine | Quarantine | case study | feasibility | qualitative | 7409 | poor | Quarantine was feasible to implement. |
| Liu et al., 2019 | United States of America | natural disaster | internally displaced | camp or camp-like | air-borne | influenza a | environmental | cleaning, disinfection, & waste management | Environmental cleaning | case study | feasibility | qualitative | 7409 | poor | Cleaning was feasible to implement. |
| Liu et al., 2019 | United States of America | natural disaster | internally displaced | camp or camp-like | air-borne | influenza a | personal protective measures | hand hygiene | Hand hygiene | case study | feasibility | qualitative | 7409 | poor | Hand hygiene measures were feasible to implement. |
| Masumbuko Claude et al., 2018 | Democratic Republic of the Congo | conflict | non-displacement | non-displacement | blood-borne | ebola | community | physical distancing | KAP survey about attitutes regarding unsafe practices | cross-sectional survey | acceptability | quantitative | 582 | poor | Some participants did not agree with Ebola-related physical distancing and case isolation measures -- 8% would touch a suspected Ebola-positive corpse and 17% would hide a suspected Ebola-positive family member from health authorities. |
| Masumbuko Claude et al., 2019 | Democratic Republic of the Congo | conflict | non-displacement | non-displacement | blood-borne | ebola | community | physical distancing | Safe and dignified burials | cross-sectional survey | acceptability | qualitative | 630 | fair | Burial practices that excluded touching the body were considered culturally unacceptable. |
| Masumbuko Claude et al., 2019 | Democratic Republic of the Congo | conflict | non-displacement | non-displacement | blood-borne | ebola | disease control | case isolation | Isolation of EVD cases | cross-sectional survey | acceptability | qualitative | 630 | fair | Isolating cases was considered unacceptable by some for fear that separation was permanent. |
| Masumbuko Claude & Hawkes, 2020 | Democratic Republic of the Congo | conflict | non-displacement | non-displacement | blood-borne | ebola | community | risk communication | Risk communication and community engagement: medical students recruited to deliver health messages | cross-sectional survey | acceptability | quantitative | 319 | poor | Lower satisfaction among participants associated with possible markers of non-compliance. Higher satisfaction scores associated with preference for local sources of EVD information over foreign response teams. CHWs overwhelmingly agreed that medical students made positive contribution to the EVD response effort. |
| Masumbuko Claude & Hawkes, 2020 | Democratic Republic of the Congo | conflict | non-displacement | non-displacement | blood-borne | ebola | community | risk communication | Risk communication and community engagement: medical students recruited to deliver health messages | cross-sectional survey | effectiveness | quantitative | 355 | poor | About half (53%) of community educators perceived that community had understood their messages. |
| Masur et al., 2017 | Haiti | informal housing | informally housed | informal housing | air-borne | tuberculosis | disease control | active case finding | Door-to-door ACF and referral to health facility of inviduals with persistent cough. | cross-sectional study | effectiveness | quantitative | 7500 | fair | Intervention led to detection of high burden of undiagnosed TB. Combined ACF and CT was an effective strategy |
| Masur et al., 2017 | Haiti | informal housing | informally housed | informal housing | air-borne | tuberculosis | disease control | contact tracing | Contact tracing undertaken for confirmed cases. | cross-sectional study | effectiveness | quantitative | 7500 | fair | Intervention led to detection of high burden of undiagnosed TB. Combined ACF and CT was an effective strategy |
| Mayaud, 2001 | United Republic of Tanzania | conflict | refugee | camp or camp-like | sexually-transmitted | hiv/aids | community | risk communication | Mass education campaigns and distribution of educational materials. Peer education and condom-use promotion among bar and brothel workers. | pre-post study | effectiveness | quantitative | 628 | poor | Health education did not impact sexual behaviour. STI prevalence increased markedly during intervention period. |
| Mayaud, 2001 | United Republic of Tanzania | conflict | refugee | camp or camp-like | sexually-transmitted | hiv/aids | personal protective measures | condom distribution | Condom distribution at clinics, during campaigns and by peer educators. | pre-post study | effectiveness | quantitative | 628 | poor | Condom distribution did not impact sexual behaviour. STI prevalence increased markedly during intervention period. |
| McGinn & Allen, 2006 | Guinea | conflict | refugee | camp or camp-like | sexually-transmitted | stis | community | risk communication | Provision of literacy classes focused on safe motherhood, family planning, STIs/HIV/AIDS and gender-based violence | cross-sectional survey | effectiveness | quantitative | 549 | fair | Intervention increased participants' evaluation of their sexual behaviour. Participants also found to retain knowledge gained in literacy classes. |
| World Health Organization, 2005 | Indonesia | natural disaster | internally displaced; non-displacement | camp or camp-like; hosted; non-displacement | various | various | community | risk communication | Health education. | case study | effectiveness | qualitative | 184864 | poor | Despite large number of displaced people, no major outbreaks occurred during the acute phase of the crisis |
| World Health Organization, 2005 | Indonesia | natural disaster | internally displaced; non-displacement | camp or camp-like; hosted; non-displacement | various | various | community | WASH | Improved WASH conditions. | case study | effectiveness | qualitative | 184864 | poor | Despite large number of displaced people, no major outbreaks occurred during the acute phase of the crisis |
| World Health Organization, 2005 | Indonesia | natural disaster | internally displaced; non-displacement | camp or camp-like; hosted; non-displacement | various | various | disease control | contact tracing | Contact tracing and follow-up. | case study | effectiveness | qualitative | 184864 | poor | Despite large number of displaced people, no major outbreaks occurred during the acute phase of the crisis |
| World Health Organization, 2005 | Indonesia | natural disaster | internally displaced; non-displacement | camp or camp-like; hosted; non-displacement | various | various | personal protective measures | hand hygiene | Distribution of soap and hygiene kits. | case study | effectiveness | qualitative | 184864 | poor | Despite large number of displaced people, no major outbreaks occurred during the acute phase of the crisis |
| Mitchell et al., 2018 | Thailand | conflict | refugee | camp or camp-like | vehicle-borne | intestinal parasites | disease control | active case finding | Pre-departure screening and treatment of refugees | pre-post study | effectiveness | quantitative | 2004 | good | Helminth infections and associated moderate-to-severe anemia decreased substantially. |
| Mobula et al., 2020 | Democratic Republic of the Congo | conflict | non-displacement | non-displacement | blood-borne | ebola | community | risk communication | Risk communication and community engagement | case study | effectiveness | qualitative | Not stated | poor | Early communication in inhabitants' local languages was helpful in establishing communication. Suitable feedback mechanisms between different sectors of the response were important. |
| Mobula et al., 2020 | Democratic Republic of the Congo | conflict | non-displacement | non-displacement | blood-borne | ebola | disease control | active case finding | Community based surveillance | case study | effectiveness | qualitative | Not stated | poor | Community involvement and effective co-ordination between different response agencies was essential. |
| Moll et al., 2007 | Honduras, Nicaragua, El Salvador & Guatemala | natural disaster | non-displacement | informal housing | water-borne | diarrhoea | community | risk communication | Community education in basic sanitation and hygiene practices | pre-post study | effectiveness | quantitative | 800 HHs | good | Integrated WASH and hygiene education programme led to improvements in access to sanitation and household water quality, and improved hand washing behaviour, associated with reduced diarrhoea in children under 3 years of age. |
| Moll et al., 2007 | Honduras, Nicaragua, El Salvador & Guatemala | natural disaster | non-displacement | informal housing | water-borne | diarrhoea | community | WASH | Provision of sustainable access to water and sanitation services | pre-post study | effectiveness | quantitative | 800 HHs | good | Integrated WASH and hygiene education programme led to improvements in access to sanitation and household water quality, and improved hand washing behaviour, associated with reduced diarrhoea in children under 3 years of age. |
| Moll et al., 2007 | Honduras, Nicaragua, El Salvador & Guatemala | natural disaster | non-displacement | informal housing | water-borne | diarrhoea | community | WASH | Provision of sustainable access to water and sanitation services | pre-post study | feasibility | quantitative | 800 HHs | good | Longer term sustainability of intervention in doubt, and chlorine disinfection not possible in all communities due to problems with supply and delivery |
| Ngwa et al., 2020 | Nigeria | conflict | internally displaced | camp or camp-like | water-borne | cholera | community | risk communication | Risk communication and raising awareness. | cross-sectional study | acceptability | qualitative | 39 | fair | Low acceptability of water chlorination due to misinformation. Initial use of unfamiliar language led to community mistrust. Mobilising key trusted members of the community was necessary. |
| Ngwa et al., 2020 | Nigeria | conflict | internally displaced | camp or camp-like | water-borne | cholera | community | risk communication | Risk communication and raising awareness. | case study | effectiveness | quantitative | population-wide | poor | Hygiene promotion was associated with low outbreak CFR |
| Ngwa et al., 2020 | Nigeria | conflict | internally displaced | camp or camp-like | water-borne | cholera | community | WASH | Hygiene promotion and use of water purification tablets | cross-sectional study | acceptability | qualitative | 39 | fair | Dislike of communal latrines and distance to them. Overcrowding led to latrine collapse and queues, further reducing acceptability. Chlorine acceptance required mobilising key trusted members of the community |
| Ngwa et al., 2020 | Nigeria | conflict | internally displaced | camp or camp-like | water-borne | cholera | community | WASH | Hygiene promotion and use of water purification tablets | case study | effectiveness | quantitative | population-wide | poor | WASH intervention was associated with low outbreak CFR |
| Ngwa et al., 2020 | Nigeria | conflict | internally displaced | camp or camp-like | water-borne | cholera | community | WASH | Repair of leak from latrine into water supply | case study | feasibility | qualitative | population-wide | poor | Delayed repair of leaking latrine may have amplified outbreak. Feasibility relies on effective co-ordination between agencies and understanding of community needs. |
| Ngwa et al., 2020 | Nigeria | conflict | internally displaced | camp or camp-like | water-borne | cholera | disease control | active case finding | ACF of cholera cases | cross-sectional study | feasibility | qualitative | 39 | fair | Outbreak declaration depended on RDTs and confirmation by laboratory culture. Lack of laboratory capacity contributed to false negative culture results and delays in outbreak confirmation. |
| O’Laughlin et al., 2014 | Uganda | conflict | refugee | camp or camp-like | sexually-transmitted | hiv/aids | disease control | active case finding | Free voluntary counselling and testing offered to clinic attendees | case-control study | acceptability | quantitative | 64,000 | good | HIV testing was acceptable. |
| O’Laughlin et al., 2014 | Uganda | conflict | refugee | camp or camp-like | sexually-transmitted | hiv/aids | disease control | active case finding | Free voluntary counselling and testing offered to clinic attendees | case-control study | effectiveness | quantitative | 64,000 | fair | The intervention resulted in more refugees screened and more cases detected. Additional side-effect was greater uptake and detection among host population |
| O’Laughlin et al., 2014 | Uganda | conflict | refugee | camp or camp-like | sexually-transmitted | hiv/aids | disease control | active case finding | Free voluntary counselling and testing offered to clinic attendees | case-control study | feasibility | quantitative | 64,000 | good | Increased testing required additional staff capacity. |
| O’Laughlin et al., 2018 | Uganda | conflict | refugee | camp or camp-like | sexually-transmitted | hiv/aids | disease control | active case finding | Screening and community engagement: home-based HIV testing | cross-sectional survey | acceptability | quantitative | 378 | fair | Home-based HIV testing was acceptable (75% accepted home-based HIV test). Odds of acceptability increased 50% per additional person in HH. Awareness campaigns about upcoming home-based testing and requests to be home at specific times may facilitate uptake. |
| O’Laughlin et al., 2018 | Uganda | conflict | refugee | camp or camp-like | sexually-transmitted | hiv/aids | disease control | active case finding | Screening and community engagement: home-based HIV testing | cross-sectional survey | feasibility | quantitative | 378 | fair | Home-based HIV testing was feasible (90% of eligible individuals encountered within 3 visits) |
| Oeltmann et al., 2008 | Thailand | conflict | refugee | camp or camp-like | air-borne | tuberculosis | disease control | active case finding | Enhanced pre-immigration screening. | case-study | effectiveness | quantitative | 15455 | poor | Intervention detected 272 cases among 15,455 people screened. |
| Okware et al., 2015 | Uganda | conflict | internally displaced | camp or camp-like | blood-borne | ebola | disease control | case isolation | Case detection, isolation, and treatment | case study | effectiveness | qualitative | Not stated | poor | Early detection of Ebola in community facilitates earlier implementation of additional control measures, frequently leading to shorter outbreak |
| Okware et al., 2015 | Uganda | conflict | internally displaced | camp or camp-like | blood-borne | ebola | disease control | quarantine | Community quarantine | case study | effectiveness | qualitative | Not stated | poor | Community enforcement was vital to successful quarantine |
| Palmer et al., 2014 | South Sudan | conflict | internally displaced | not specified | vector-borne | human african trypanosomiasis | disease control | active case finding | ACF for HAT | case study | acceptability | qualitative | 114 | poor | Open-door policy to screening empowered displaced Dinka to use HAT services more actively, leading to higher service uptake compared to Madi returnees |
| Phillips et al., 2015 | South Sudan | conflict | refugee | camp or camp-like | water-borne | hepatitis A, hepatitis E, diarrhoea | community | risk communication | Hand hygiene education campaign | cross-sectional survey | effectiveness | quantitative | 600 | fair | Despite high levels of message comprehension, practices were unchanged. Majority washed hands, but only minority used soap. |
| Plummer, 1995 | United Republic of Tanzania | conflict | refugee | camp or camp-like | water-borne | cholera | community | risk communication | CHWs targeted at-risk groups with health education campaign about hand washing, and trained community members to do the same. | case study | effectiveness | qualitative | Not stated | poor | Education and active case finding ensured most cholera patients arrived in stable condition, improving chances of survivial |
| Plummer, 1995 | United Republic of Tanzania | conflict | refugee | camp or camp-like | multiple | multiple | disease control | active case finding | CHWs trained in active case finding for patients with various conditions. | case study | effectiveness | qualitative | Not stated | poor | Active case finding and referral were described as 'instrumental in lowering morbidity and mortality rates' |
| Plummer, 1995 | United Republic of Tanzania | conflict | refugee | camp or camp-like | water-borne | cholera | disease control | active case finding | During the cholera epidemic, CHWs focused on mass education and active case finding. | case study | effectiveness | qualitative | Not stated | poor | Education and active case finding ensured most cholera patients arrived in stable condition, improving chances of survivial |
| Protopopoff et al., 2007 | Burundi | conflict | non-displacement | non-displacement | vector-borne | malaria | environmental | vector control | Implementation of IRS and bednet distribution during malaria epidemic | controlled intervention study | feasibility | quantitative | 340 HHs | poor | Anopheles indoor resting reduced, but malaria incidence unchanged with intervention |
| Protopopoff et al., 2007 | Burundi | conflict | non-displacement | non-displacement | vector-borne | malaria | personal protective measures | vector protection | Implementation of IRS and bednet distribution during malaria epidemic | controlled intervention study | feasibility | quantitative | 340 HHs | poor | Anopheles indoor resting reduced, but malaria incidence unchanged with intervention |
| Reif et al., 2016 | Haiti | informal housing | informally housed | informal housing | sexually-transmitted | hiv/aids, syphilis, gonorrhea, chlamydia | disease control | active case finding | ACF for TB and a variety of STIs. Package of approaches included community sensitization by community HCWs and targeting high-risk groups. | cross-sectional study | acceptability | quantitative | 3425 | good | ACF was highly acceptable. Testing bundled with screening for other conditions, e.g. TB, lowering stigma associated with HIV. Testing was 'opt-out' rather than 'opt-in'. Screening done in community setting, increasing access to teens. |
| Richards et al., 2009 | Myanmar | conflict | internally displaced | camp or camp-like | vector-borne | malaria | community | risk communication | Community sensitization by educational malaria messages addressing causes, importance of early treatment seeking, adherence to medicines, and use of LLITNs. | pre-post study | effectiveness | quantitative | 712 | good | Increased knowledge about malaria symptoms, preventative measures, and available treatments |
| Richards et al., 2009 | Myanmar | conflict | internally displaced | camp or camp-like | vector-borne | malaria | community | risk communication | Provision of educational messages. | pre-post study | feasibility | quantitative | 3431 | fair | Population reached by programme almost doubled in 18 months (from 1868 to 3431) |
| Richards et al., 2009 | Myanmar | conflict | internally displaced | camp or camp-like | vector-borne | malaria | disease control | active case finding | Early diagnosis and treatment | pre-post study | effectiveness | quantitative | 3431 | good | P. falciparum prevalence and incidence declined |
| Richards et al., 2009 | Myanmar | conflict | internally displaced | camp or camp-like | vector-borne | malaria | personal protective measures | vector protection | Free distribution of LLITNs | pre-post study | acceptability | quantitative | 3431 | good | LLITN usage was highly acceptable, increasing from 0% to ~90% |
| Richards et al., 2009 | Myanmar | conflict | internally displaced | camp or camp-like | vector-borne | malaria | personal protective measures | vector protection | Free distribution of LLITNs | pre-post study | effectiveness | quantitative | 3431 | good | P. falciparum prevalence and incidence declined |
| Richards et al., 2009 | Myanmar | conflict | internally displaced | camp or camp-like | vector-borne | malaria | personal protective measures | vector protection | Free distribution of LLITNs | pre-post study | feasibility | quantitative | 3431 | fair | Population reached by programme almost doubled in 18 months (from 1868 to 3431) |
| Roberts et al., 2001 | Malawi | conflict | refugee | camp or camp-like | water-borne | diarrhoeal | personal protective measures | water purification | Distribution of water containers with improved design | controlled intervention study | effectiveness | qualitative | 400 HHs | poor | Diarrhoea incidence was non-significantly recued among intervention HHs compared to control |
| Rowland et al., 1994 | Pakistan | conflict | refugee | camp or camp-like | vector-borne | malaria | environmental | vector control | Indoor residual spraying of two insecticides (malathion to lambdacyhalothrin) | pre-post study | effectiveness | quantitative | ~27500 | good | High IRS coverage achieved (>85%). Spraying with lambdacyhalothrin effectively reduced P. falciparum prevalence whilst malathion did not. Neither insecticide effectively reduced P. vivax infections. Spraying was more effective in summer than in spring. |
| Rowland et al., 1996 | Pakistan | conflict | refugee | informal housing | vector-borne | malaria | personal protective measures | vector protection | Distribution of permethrin-impregnated bednets, with education on use. | controlled intervention study | effectiveness | quantitative | 2792 | fair | Bednets were effective against malaria |
| Rowland, Hewitt, Durrani, Bano, et al., 1997 | Pakistan | conflict | refugee | camp or camp-like | vector-borne | malaria | environmental | vector control | Indoor residual spraying of insecticide | pre-post study | effectiveness | quantitative | 14 villages | fair | Villages sprayed in July had 62% reduced incidence compared with 15% in April. |
| Rowland, Hewitt, Durrani, Bano, et al., 1997 | Pakistan | conflict | refugee | camp or camp-like | vector-borne | malaria | environmental | vector control | Indoor residual spraying of insecticide | pre-post study | feasibility | quantitative | 14 villages | fair | High coverage of IRS was feasible. |
| Rowland, Hewitt, Durrani, Saleh, et al., 1997 | Pakistan | conflict | refugee | camp or camp-like | vector-borne | malaria | personal protective measures | vector protection | Pyrethroid-impregnated bednets distributed, re-impregnated, and sold | controlled intervention study | effectiveness | quantitative | 359 HHs | fair | Bednet usage significantly reduced malaria prevalence |
| Rowland et al., 1999 | Pakistan | conflict | refugee | camp or camp-like | vector-borne | malaria | personal protective measures | vector protection | Distribution of permethrin-treated headscarfs and top-sheets | controlled intervention study | effectiveness | quantitative | 825 | fair | Intervention was effective at reducing malaria incidence |
| Rowland et al., 2004 | Pakistan | conflict | refugee | camp or camp-like | vector-borne | malaria | personal protective measures | vector protection | Distribution of Mosbar mosquito repellent | controlled intervention study | effectiveness | quantitative | 1148 | good | Repellent was effective against malaria |
| Ruckstuhl et al., 2017 | Central African Republic | conflict | internally displaced | hosted | vector-borne | malaria | disease control | active case finding | ACF by CHWs to improve access to malaria care | case-study | feasibility | quantitative | 80 villages | fair | Intervention was feasible: 198,382 people consulted CHW; 81% were malaria positive by rapid diagnostic test. 98.9% of cases received artemisinin-based combination therapy. |
| Samal & Dehury, 2017 | India | informal housing | non-displacement | informal housing | air-borne | tuberculosis | community | risk communication | Awareness campaign to inform households about TB symptoms, diagnosis, treatment and associated services | pre-post study | effectiveness | quantitative | 100 HHs | poor | Intervention led to substantial change in health-seeking behaviour from private to publicly provided. |
| Scobie et al., 2016 | Thailand | conflict | refugee | camp or camp-like | water-borne | cholera | community | risk communication | Cholera education campaign | cross-sectional survey | effectiveness | quantitative | 271 | fair | Intervention led to increased awareness of cholera prevention strategies. |
| Scobie et al., 2016 | Thailand | conflict | refugee | camp or camp-like | water-borne | cholera | community | WASH | Distribution of WASH ietms | cross-sectional survey | effectiveness | quantitative | 271 | fair | Reported receipt of chlorine solution and water containers decreased while receipt of soap was unchanged. |
| Sekine & Roskosky, 2018 | Nepal | natural disaster | internally displaced | camp or camp-like | water-borne | cholera | community | risk communication | Hygiene promotion and awareness-raising campaigns by CHWs at cholera hotspots | case study | effectiveness | qualitative | Not stated | poor | Safe water purification and storage remained low. |
| Sekine & Roskosky, 2018 | Nepal | natural disaster | internally displaced | camp or camp-like | water-borne | cholera | community | WASH | Water treatment product distribution and water quality testing | case study | feasibility | qualitative | Not stated | poor | Challenges to feasibility included lack of fast-track funding and poor co-ordination between health and WASH sectors, leading to delays in response and confusion regarding implementation. |
| Shortus et al., 2016 | Solomon Islands | natural disaster | internally displaced | camp or camp-like | vector-borne | dengue | disease control | active case finding | Implementation of a sentinel-site EWARS | pre-post | effectiveness | qualitative | ~10,000 | poor | The system identified a malaria outbreak |
| Shortus et al., 2016 | Solomon Islands | natural disaster | internally displaced | camp or camp-like | vector-borne | dengue | environmental | vector control | LLITNs, IRS, and larviciding conducted in high risk areas | pre-post | effectiveness | qualitative | ~10,000 | poor | Vector control measures likely prevented an increase in vector-borne disease, but lack of disease surveillance and entomological monitoring prevented formal assessment |
| Shortus et al., 2016 | Solomon Islands | natural disaster | internally displaced | camp or camp-like | vector-borne | dengue | environmental | vector control | LLITNs, IRS, and larviciding conducted in high risk areas | pre-post | feasibility | qualitative | ~10,000 | poor | Despite high coverage, data availability and logistical issues impacted feasibility |
| Shortus et al., 2016 | Solomon Islands | natural disaster | internally displaced | camp or camp-like | vector-borne | dengue | personal protective measures | vector protection | LLITNs, IRS, and larviciding of Aedes breeding sites conducted in high risk areas | pre-post | feasibility | quantitative | ~10,000 | poor | Intervention was feasible: 80% LLITN, and 77% IRS and larviciding coverage, acheived in first week |
| Sircar et al., 1987 | India | informal housing | informally housed | informal housing | water-borne | diarrhoeal | personal protective measures | hand hygiene | Handwashing promotion with soap distribution | controlled intervention study | effectiveness | quantitative | 3668 | poor | Handwashing was effective against dysentry for those over 5 years but not those under 5. Not effective against watery diarrhoea for any age group. |
| Soares et al., 2013 | Brazil | informal housing | informally housed | informal housing | air-borne | tuberculosis | community | risk communication | CHWs conducted risk communication (TB awareness sessions and establishing community support network), DOTS, ACF and contact tracing. | pre-post study | effectiveness | quantitative | ~60000 | good | The interventions improved treatment and decreased defaulting. Case rates declined, but not statistically significantly. |
| Soares et al., 2013 | Brazil | informal housing | informally housed | informal housing | air-borne | tuberculosis | disease control | active case finding | CHWs conducted risk communication (TB awareness sessions and establishing community support network), DOTS, ACF and contact tracing. | pre-post study | effectiveness | quantitative | ~60000 | good | The interventions improved treatment and decreased defaulting. Case rates declined, but not statistically significantly. |
| Soares et al., 2013 | Brazil | informal housing | informally housed | informal housing | air-borne | tuberculosis | disease control | contact tracing | CHWs conducted risk communication (TB awareness sessions and establishing community support network), DOTS, ACF and contact tracing. | pre-post study | effectiveness | quantitative | ~60000 | good | The interventions improved treatment and decreased defaulting. Case rates declined, but not statistically significantly. |
| Spencer et al., 2004 | Uganda | conflict | internally displaced | camp or camp-like | vector-borne | malaria | community | risk communication | Education campaign on ITN use in schools and camps | cross-sectional survey | effectiveness | quantitative | 3298 | fair | Intervention was possibly effective at changing ITN storage behaviour |
| Spencer et al., 2004 | Uganda | conflict | internally displaced | camp or camp-like | vector-borne | malaria | personal protective measures | vector protection | HH distribution of ITN | cross-sectional survey | effectiveness | quantitative | 3298 | fair | ITN users had lower risk of malaria than non-users |
| Sundnes & Haimanot, 1993 | Ethiopia | conflict | prisoners of war | camp or camp-like | vector-borne | louse-borne relapsing fever | environmental | vector control | Delousing | controlled intervention study | effectiveness | quantitative | Not stated | Poor | Effective control of louse-borne relapsing fever epidemic was dependent on efficient vector control in addition to antibiotic treatment. |
| Takahashi et al., 2013 | Japan | natural disaster | internally displaced | camp or camp-like | air-borne | measles | community | risk communication | Infected volunteers were asked refrain from working in shelters | case study | effectiveness | quantitative | Not stated | poor | Messaging increased number of symptomatic volunteers staying away from post-earthquake shelters |
| Tanaka et al., 2008 | United Republic of Tanzania | conflict | refugee | camp or camp-like | sexually-transmitted | hiv/aids | community | risk communication | Youth peer education and Voluntary Counselling and Testing services. Reproductive education followed by serological testing. Condom distribution. | cross-sectional survey | effectiveness | quantitative | 1140 | poor | Knowledge of HIV/AIDS and its prevention was generally high, but practice of HIV risk reduction was low. High levels of non-regular sexual partners. Poorer knowledge, attitudes and practices among females. |
| Tappero & Tauxe, 2011 | Haiti | natural disaster | non-displacement, internally displaced | non-displacement, camp or camp-like | water-borne | cholera | community | risk communication | CHWs provided health education and led community-level prevention. | case study | feasibility | qualitative | Not stated | poor | Additional funding was required to ensure CHWs received appropriate training |
| Tappero & Tauxe, 2011 | Haiti | natural disaster | non-displacement, internally displaced | non-displacement | water-borne | cholera | community | WASH | Distribution of water purification tablets and enhanced chlorination of water | case study | feasibility | qualitative | Not stated | poor | Intervention was successfully implemented by multiple organisations |
| Tappero & Tauxe, 2011 | Haiti | natural disaster | non-displacement, internally displaced | non-displacement | water-borne | cholera | disease control | active case finding | Increased diagnostic laboratory testing capacity | case study | feasibility | qualitative | Not stated | poor | Intervention was feasible and enabled rapid confirmation of cholera outbreak |
| Thomson et al., 2013 | South Sudan | conflict | refugee | camp or camp-like | water-borne | hepatitis e | community | risk communication | HEV preventive hygiene education conducted during HH visits, at health facilities, and in community forums. | case study | feasibility | qualitative | Not stated | poor | Risk communication measures were feasible to implement |
| Thomson et al., 2013 | South Sudan | conflict | refugee | camp or camp-like | water-borne | hepatitis e | community | WASH | Increased availability of treated drinking water, increased latrine coverage, distribution of soap and water storage vessels, installation of handwashing stations, and expanded hygiene promotion activities | case study | effectiveness | qualitative | Not stated | poor | Targeted WASH measures alone were not effective at rapidly reducing Hep. E incidence |
| Thomson et al., 2013 | South Sudan | conflict | refugee | camp or camp-like | water-borne | hepatitis e | community | WASH | Increased availability of treated drinking water, increased latrine coverage, distribution of soap and water storage vessels, installation of handwashing stations, and expanded hygiene promotion activities | case study | feasibility | qualitative | Not stated | poor | WASH interventions were feasible to implement |
| Thomson et al., 2013 | South Sudan | conflict | refugee | camp or camp-like | water-borne | hepatitis e | disease control | active case finding | Active surveillance implemented by training CHWs to detect and refer jaundiced patients | case study | feasibility | qualitative | Not stated | poor | ACF by CHWs was feasible in this setting |
| Tong et al., 2011 | Democratic Republic of the Congo | conflict | non-displacement; internally displaced | not specified | vector-borne | human african trypanosomiasis | disease control | active case finding | ACF by mobile health teams | case study | feasibility | qualitative | Not stated | poor | Widespread screening of rural populations was feasible, but insecurity led to disruptions resulting in reduced geographic covered. |
| Toscani & Richard, 1988 | Sudan | food crisis | refugee | camp or camp-like | air-borne | tuberculosis | disease control | active case finding | Refugee population trained as medical assistants and home-visitors; HHs visited daily for screening. Establishment of laboratory for diagnostic testing. | case study | acceptability | qualitative | 6250 | poor | Low acceptability of screening for fear of consequences of positive test |
| Toscani & Richard, 1988 | Sudan | food crisis | refugee | camp or camp-like | air-borne | tuberculosis | disease control | active case finding | Refugee population trained as medical assistants and home-visitors; HHs visited daily for screening. Establishment of laboratory for diagnostic testing. | case study | feasibility | quantitative | 6250 | poor | Intervention across entire camp was feasible, but issues with test sensitivity and follow up led to various treatment difficulties. Programme was interrupted by return of refugees. |
| Vinck et al., 2019 | Democratic Republic of the Congo | conflict | non-displacement | non-displacement | blood-borne | ebola | community | risk communication | RCCE intervention providing EVD messaging regarding safe practices | cross-sectional study | effectiveness | quantitative | 961 | good | Widespread belief in misinformation. Low community trust of the response. Low social interaction (~3%) and public space (~20%) avoidance, moderate physical contact avoidance (~50%), high direct avoidance (~80%) of suspect EVD cases. |
| Walden et al., 2005 | Sudan | conflict | internally displaced | camp or camp-like | water-borne | shigellosis | environmental | cleaning, disinfection, & waste management | Mass disinfection of water containers with chlorine | pre-post study | effectiveness | qualitative | 6900 HHs | fair | Though diarrhoea outbreak had peaked prior to intervention, incidence continued to decrease afterwards |
| Walden et al., 2005 | Sudan | conflict | internally displaced | camp or camp-like | water-borne | shigellosis | environmental | cleaning, disinfection, & waste management | Mass disinfection of water containers with chlorine | pre-post study | feasibility | quantitative | 6900 HHs | fair | Intervention was a feasible and rapid intervention (88% of containers disinfected in 5 days). |
| Woodward et al., 2011 | Guinea | conflict | refugee | camp or camp-like | sexually-transmitted | hiv/aids | community | risk communication | Refugee-led peer health education. | cross-sectional study | effectiveness | quantitative | 889 | good | Intervention led to improved HIV attitudes and protective behaviours |
| Woodward et al., 2011 | Guinea | conflict | refugee | camp or camp-like | sexually-transmitted | hiv/aids | personal protective measures | condom distribution | Free condom distribution | cross-sectional study | feasibility | qualitative | 889 | good | Condom distribution was feasible and acceptable, but supply did not always meet demand. Challenges for women as men decided condom use. Male-targeted condom promotion campaign could increase uptake. |
| HeRAMS Summary Report. Syrian Arab Republic. Quarter 1, 2015, 2015 | Syria | conflict | not specified | not specified | not specified | not specified | disease control | active case finding | Availability of laboratory capacity | cross-sectional survey | feasibility | quantitative | 1777 HFs | poor | 44% HFs had basic laboratory capacity |
| Yamada et al., 2006 | Sri Lanka | natural disaster | internally displaced | camp or camp-like | vector-borne | multiple | disease control | active case finding | ACF with immediate isolation and transfer of cases with symptoms of diarrhea, respiratory infections, and conjunctivitis | case study | feasibility | qualitative | Not stated | poor | ACF was feasible, but response efforts were poorly co-ordinated between actors. |
| Yamada et al., 2006 | Sri Lanka | natural disaster | internally displaced | camp or camp-like | vector-borne | malaria, dengue | environmental | vector control | Fogging around displacement camps | case study | feasibility | qualitative | Not stated | poor | No outbreaks occurred during post-disaster acute phase after fogging |
| Yeasmin et al., 2017 | Bangladesh | informal housing | informally housed | informal housing | water-borne | not specified | community | risk communication | Behavioural change messaging to encourage appropriate waste disposal. | pre-post study | effectiveness | qualitative | 8 | poor | Intervention led to reduced toilet blockage |
| Yeasmin et al., 2017 | Bangladesh | informal housing | informally housed | informal housing | water-borne | not specified | community | WASH | Provision of waste bins with lids in communal latrines | cross-sectional study | acceptability | qualitative | 8 | poor | Bins were easy-to-use and improvements in appearance of toilet and surrounding environment were reported. However, acceptability affected by feelings of disgust |
| Yeasmin et al., 2017 | Bangladesh | informal housing | informally housed | informal housing | water-borne | not specified | community | WASH | Provision of waste bins with lids in communal latrines | pre-post study | effectiveness | qualitative | 8 | poor | Intervention led to reduced toilet blockage |
| Zeng, 2008 | China | natural disaster | internally displaced | camp or camp-like | water-borne | multiple | community | WASH | Provision of toilets and bottled water | case study | feasibility | qualitative | Not stated | poor | Feasible to construct toilets but not maintain them; Feasible to distribute bottled water for drinking, but only in the short term |
| Zhao et al., 2018 | Myanmar | informal housing | internally displaced | camp or camp-like | vector-borne | malaria | environmental | vector control | Indoor residual spraying | cohort study | effectiveness | quantitative | 1680 | poor | Risk of malaria significantly lower among those using IRS |
| Zhao et al., 2018 | Myanmar | informal housing | internally displaced | camp or camp-like | vector-borne | malaria | personal protective measures | vector protection | Provision of bednets | cohort study | effectiveness | quantitative | 1680 | poor | Risk of malaria significantly lower among those using bednets |
